# Lipidomic profiling of human serum enables detection of pancreatic cancer

**DOI:** 10.1101/2021.01.22.21249767

**Authors:** Denise Wolrab, Robert Jirásko, Eva Cífková, Marcus Höring, Ding Mei, Michaela Chocholoušková, Ondřej Peterka, Jakub Idkowiak, Tereza Hrnčiarová, Ladislav Kuchař, Robert Ahrends, Radana Brumarová, David Friedecký, Gabriel Vivo-Truyols, Pavel Škrha, Jan Škrha, Radek Kučera, Bohuslav Melichar, Gerhard Liebisch, Ralph Burkhardt, Markus R. Wenk, Amaury Cazenave-Gassiot, Petr Karásek, Ivo Novotný, Roman Hrstka, Michal Holčapek

## Abstract

Pancreatic cancer has the worst prognosis among all cancers^1^. Cancer screening programs based on the analysis of body fluids can improve the survival time of patients, who are often diagnosed too late at an incurable stage^2^. Several studies have reported the dysregulation of lipid metabolism in tumor cells and tissues^3^, suggesting that the changes of blood lipidome may accompany tumor growth and progression. Analytical methods based on mass spectrometry (MS) using either direct infusion or chromatographic separation^4^ are convenient for high-throughput lipidomic profiling. Here we show that the comprehensive quantitation of a wide range of serum lipids reveals statistically significant differences between pancreatic cancer patients and healthy controls visualized by multivariate data analysis. Initial results for 364 human serum samples in the discovery phase were subsequently verified in the qualification phase on 554 samples measured by three independent laboratories, and finally on 830 samples from four blood collection sites in the verification phase. Concentrations suggestive of dysregulation of some very long chain sphingomyelins (SM 42:1, SM 41:1, SM 39:1, and SM 40:1), ceramides (Cer 41:1, and Cer 42:1), and (lyso)phosphatidylcholines (LPC 18:2) were recorded. Some lipid species indicated a potential as biomarkers of survival. The sensitivity and specificity to diagnose pancreatic cancer is over 90%, which outperforms CA 19-9, especially in early stage, and is comparable to established imaging diagnostic methods. The accuracy of lipidomic approach is not influenced by the cancer stage, analytical method, or blood collection site.

---

Early cancer diagnosis based on non-invasive screening has been one of the major unmet needs in medical research over the last decades^1^. Some cancer types, such as pancreatic cancer^2^, do not show specific symptoms making the diagnosis at an early stage difficult. Pancreatic ductal adenocarcinoma (PDAC), accounting for 90% of pancreatic cancers, is mostly diagnosed at late stage resulting in the worst 5-year survival rate (7%) among all cancers^5^. Imaging modalities used to diagnose PDAC in clinical practice included magnetic resonance imaging, computed tomography, endoscopic ultrasound, and positron emission tomography, with accuracies reported in the meta-analysis of 5399 patients from 52 studies of 90%, 89%, 89%, and 84%, respectively^6^. Invasive procedures, *i*.*e*., biopsies, were performed only for the final confirmation of PDAC. Several types of blood tests were considered for PDAC screening^7-9^, such as carbohydrate antigen (CA) 19-9 measured alone or with other blood proteins, *e*.*g*., carcinoembryonic antigen. The sensitivity and specificity of CA 19-9 were around 80% for advanced stages of PDAC, but dropped to 30-50% for small non-metastatic tumors^10^, which prevents the utilization for early screening. *Kirsten-ras* (*KRAS*) mutation testing currently used in the clinical practice for epithelial cancers (*e*.*g*., lung or colorectal cancers) was considered for PDAC diagnostic using liquid biopsies, but the sensitivity was too low. This mutation encountered in more than 90% of PDAC^11^ and was related to inferior overall survival. *KRAS* may be involved in the metabolic reprogramming of fast proliferating tumor cell population towards elevated glucose and glutamine flows defined as one of the hallmarks of cancer^12^. Furthermore, the uptake of nutrients in *KRAS* mutated cells can include blood lipids for the cell proliferation and survival^13,14^. *KRAS* mutation has been reported to be associated with the lipid metabolism in pancreatic cancer cells^15^. Lipids have numerous functions in human metabolism^16^. Changes in the lipid metabolism were already reported in other cancer types^3^, mostly for cell lines^17^, tissues^18^, but less frequently for body fluids^19^. MS based lipidomic analysis has proven to be robust for high-throughput quantitation^20^ and in combination with multivariate data analysis even small differences in lipid profiles can be detected^3^.

In the present study, we investigated the potential of comprehensive lipidomic profiling of human serum for PDAC detection. In most cases, the monitoring of single lipid species did not provide a reliable differentiation between cases and controls. Lipid species and classes are interrelated, therefore a multi-analyte approach reflecting the whole lipidome provides a stronger experimental design for clinical diagnostics. The overall methodologic setup is described in Fig. 1 and in Methods. Exogenous lipid class internal standards (IS) were added to serum before liquid-liquid extraction (Supplementary Table 1). Prepared extracts were analyzed by several MS approaches in individual study phases^21^ (Fig. 1). The analytical validation of quantitative methods was performed in the Phase I in accordance with bioanalytical validation guidelines^22,23^ including two steps of quality control (details in Methods)^24^. Lipidomic MS data were processed using in-house script^17,18^, and then statistically evaluated by multivariate data analysis. The data set was split into training and validation sets. The training set was used for building statistical models, which were then applied for the prediction of samples from the validation set to verify the method performance for samples with unknown health status. The initial Phase I included 364 PDAC patients and healthy control samples, which were analyzed by ultrahigh-performance supercritical fluid chromatography (UHPSFC)/MS^24,25^, shotgun MS, and matrix-assisted laser desorption/ionization (MALDI)-MS^26^. Small differences were observed in the lipidome between males and females, therefore gender separated statistical models were further used in this work (Extended Data Fig. 1). Subsequently, an extended cohort of 554 samples was analyzed in parallel by three independent laboratories and four different MS based approaches (Phase II). Finally, 830 samples from four collection sites were analyzed by UHPSFC/MS (Phase III). The method potential for diagnostic and prognosis purposes was evaluated with advanced multivariate and univariate biostatistical tools. Molar concentrations (nmol/mL) of all quantified lipid species (Supplementary Table 2) were used for all statistical analyses and visualizations (Supplementary Tables 3 – 5). An overview of all human subjects and clinical information was provided in Supplementary Tables 6 and 7.

**Fig. 1.**
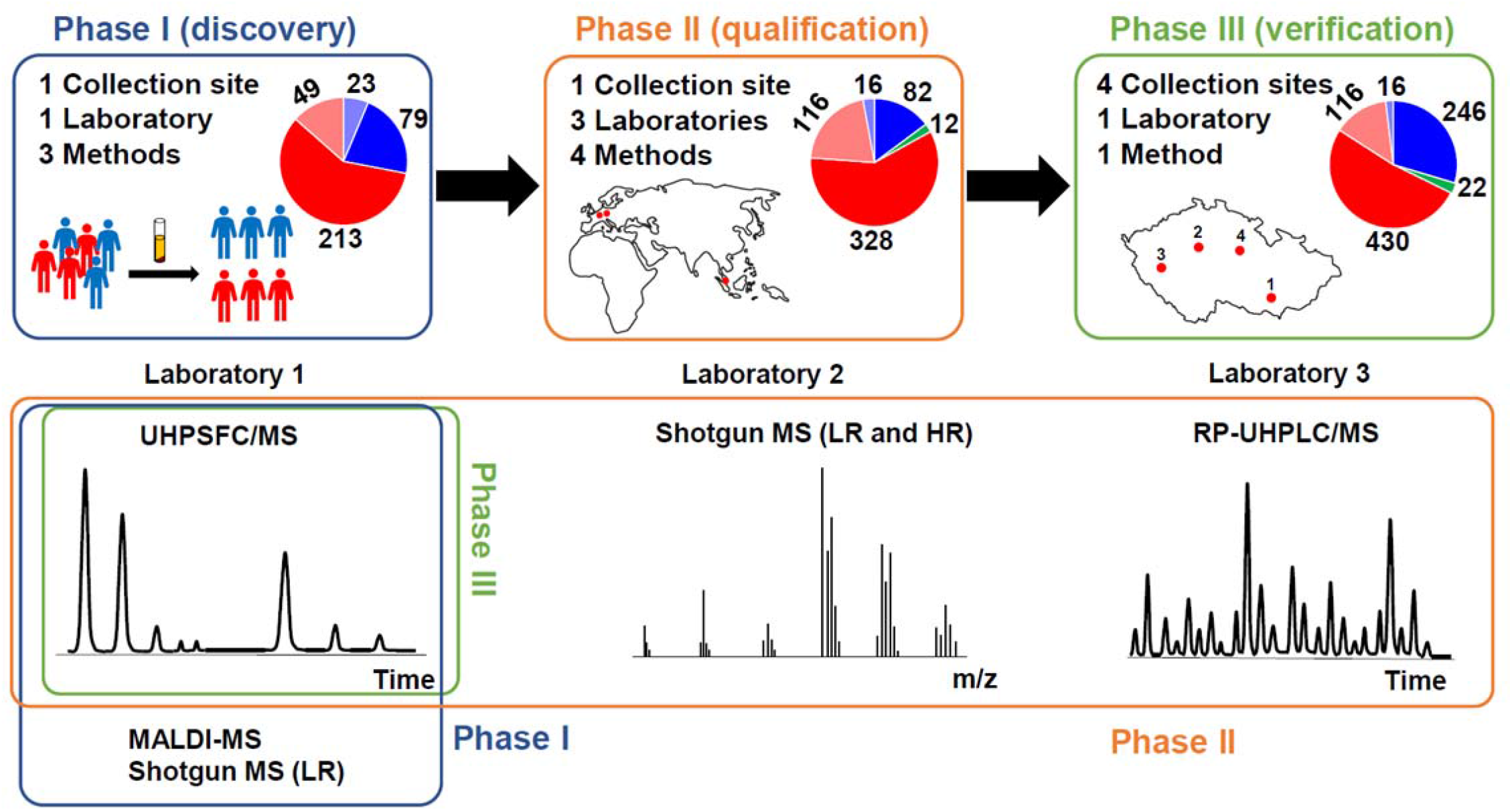
Overview of study design for the differentiation of PDAC patients (T, red) from normal healthy controls (N, blue) and pancreatitis patients (Pan, green) based on the lipidomic profiling of human serum using various mass spectrometry based approaches. **a**, Phase I (discovery) for 364 samples (262 T + 102 N) divided into training (213 T + 79 N) and validation (49 T + 23 N) sets measured by UHPSFC/MS, shotgun MS (LR), and MALDI-MS. **b**, Phase II (qualification) for 554 samples (444 T + 98 N + 12 Pan) divided into training (328 T + 82 N + 12 Pan) and validation (116 T + 16 N) sets measured by UHPSFC/MS, shotgun MS (LR and HR), and RP-UHPLC/MS at 3 different laboratories. **c**, Phase III (verification) for 830 samples (546 T + 262 N + 22 Pan) divided into training (430 T + 246 N + 22 Pan) and validation (116 T + 16 N) sets measured by UHPSFC/MS for samples obtained from 4 collection sites.

The statistical evaluation of UHPSFC/MS and shotgun MS data in the Phase I showed a partial discrimination between cases and controls in principal component analysis (PCA) score plots and distinct group differentiation when using supervised orthogonal projections to latent structures discriminant analysis (OPLS-DA) models (Extended Data Fig. 1b,c and 2a,b). The predicted response values for training and validation sets obtained from OPLS-DA models based on the training set were used for building receiver operating characteristic (ROC) curves. The area under the curve (AUC) values were over 0.99 for the training set and over 0.93 for the validation set (Extended Data Fig. 2c,d). Box plots for SM 41:1 illustrated the same trend for both methods (Extended Data Fig. 2e,f). MALDI-MS, performed on a limited number of 64 samples, provided complementary information about the down-regulation of some anionic glycosphingolipids, such as sulfatides (Extended Data Fig. 2g,h). However, MALDI-MS measurements were not continued in the next phases, as the approach is semi-quantitative.

The goal of the Phase II was to verify the results obtained in the Phase I by independent laboratories. The extended cohort of 554 samples was measured by four different MS based methods (UHPSFC/MS, shotgun MS with low-resolution (LR) and high-resolution (HR), and RP-UHPLC/MS) with different lipidomic coverage (Extended Data Fig. 3). Results from Phases II and III were normalized to reported values of lipid species concentrations^27^ in the NIST reference material according to previously published work^28^ (Supplementary Tables 4 and 5). ROC curves (Fig. 2a-d), OPLS-DA score plots, sensitivity, specificity, and accuracy prepared separately for males (Extended Data Fig. 4a-h) and females (Extended Data Fig. 5a-h) indicated a clear discrimination of case and control groups for both training and validation sets. Box plots constructed for the most significantly dysregulated lipid species (Fig. 2e-g, Extended Data Fig. 4i, 5i, and 6) revealed a mutual comparability of molar concentrations from individual laboratories, despite the use of different approaches for the sample preparation and lipidomic quantitation. Based on the Phase II results, we hypothesize that outcomes should be reproducible for other laboratories experienced in the lipidomic analysis. An important issue for PDAC screening is the differentiation between PDAC and chronic pancreatitis patients (Extended Data Fig. 4i and 5i). Although, lipid profiles of chronic pancreatitis patients are comparable to healthy controls, the number of collected blood samples was not yet sufficient to draw significant conclusions, but this should be studied for a larger number of subjects within planned clinical validation.

**Fig. 2.**
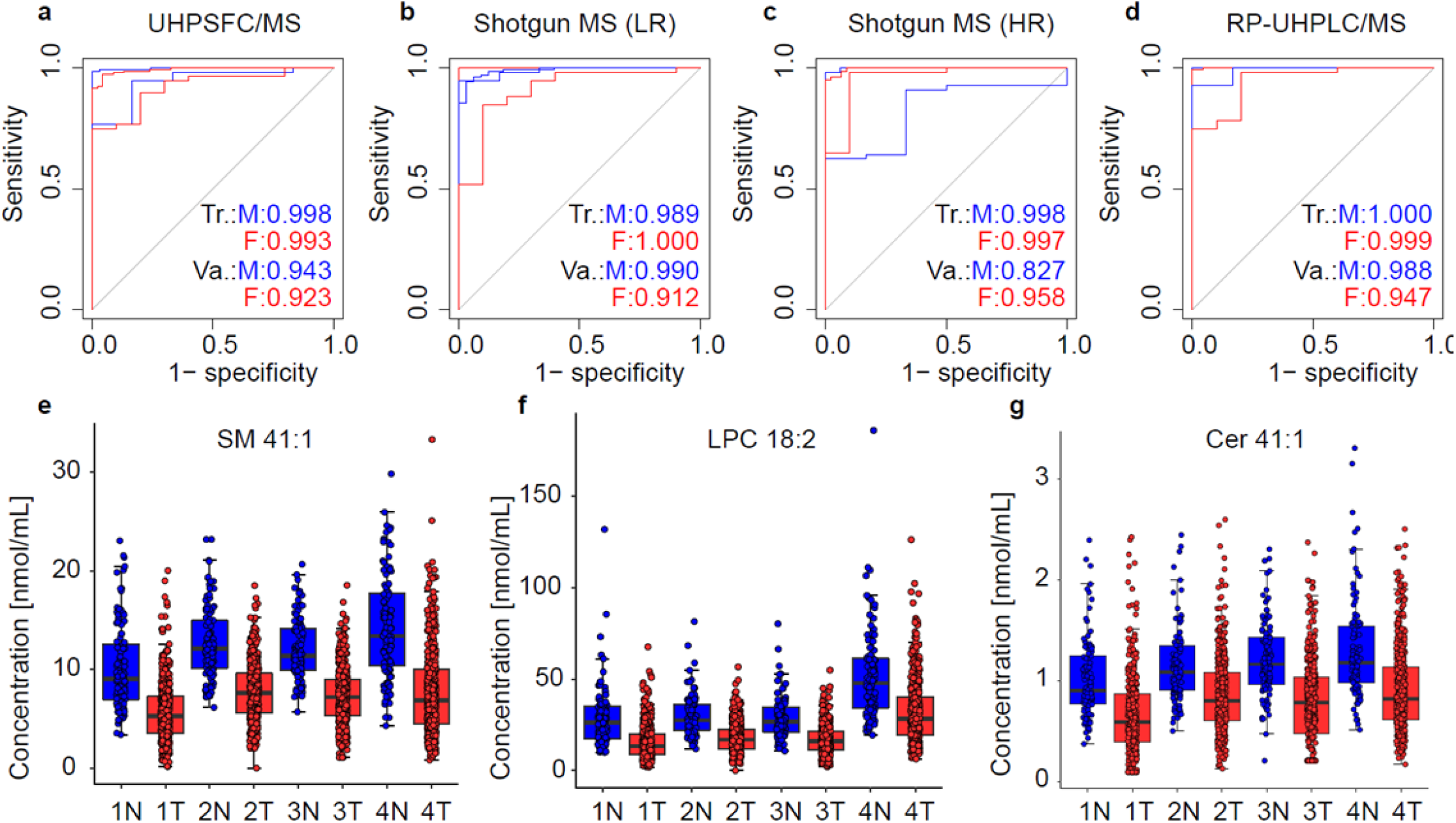
Comparison of Phase II results obtained at three different laboratories using four mass spectrometry based approaches. ROC curves for males (M) and females (F) in training (Tr.) and validation (Va.) sets: **a**, UHPSFC/MS, **b**, shotgun MS (LR), **c**, shotgun MS (HR), and **d**, RP-UHPLC/MS. Box plots of lipid concentrations normalized with the NIST reference material for samples obtained from PDAC patients (443 T) and healthy controls (95 N) of both genders including both validation and training sets: **e**, SM 41:1, **f**, LPC 18:2, and **g**, Cer 41:1 for UHPSFC/MS (Method 1), shotgun MS (LR) (Method 2), shotgun MS (HR) (Method 3), and RP-UHPLC/MS (Method 4).

In the Phase III, we investigated the method sensitivity for different blood collection sites, applicability for the early stage (T1 or T2) screening, effects of surgery, systemic therapy, and diabetes mellitus on lipidomic profiles. Statistical models for males (Fig. 3) and females (Extended Data Fig. 7 and 8a) included 830 subjects from four collections sites, before and during the treatment, before and after the surgery, without and with diabetes mellitus. PCA score plots indicated minor group differentiation (Fig. 3a and Extended Data Fig. 7a), but OPLS-DA (Fig. 3b and Extended Data Fig. 7b) captured differences between PDAC and controls, as illustrated by sensitivity, specificity, and accuracy values (Fig. 3c, Extended Data Fig. 7c, and Supplementary Table 8). Lipid species with the highest influence on group clustering were selected based on S-plots (Fig. 3d and Extended Data Fig. 7d), box plots (Extended Data Fig. 6), and statistical tests (Supplementary Table 9), and listed with their fold changes, p-values, T-values, and variable influence of projection (VIP) values. Heat maps were generated based on the most dysregulated lipids (Fig. 3e and Extended Data Fig. 7e).

**Fig. 3.**
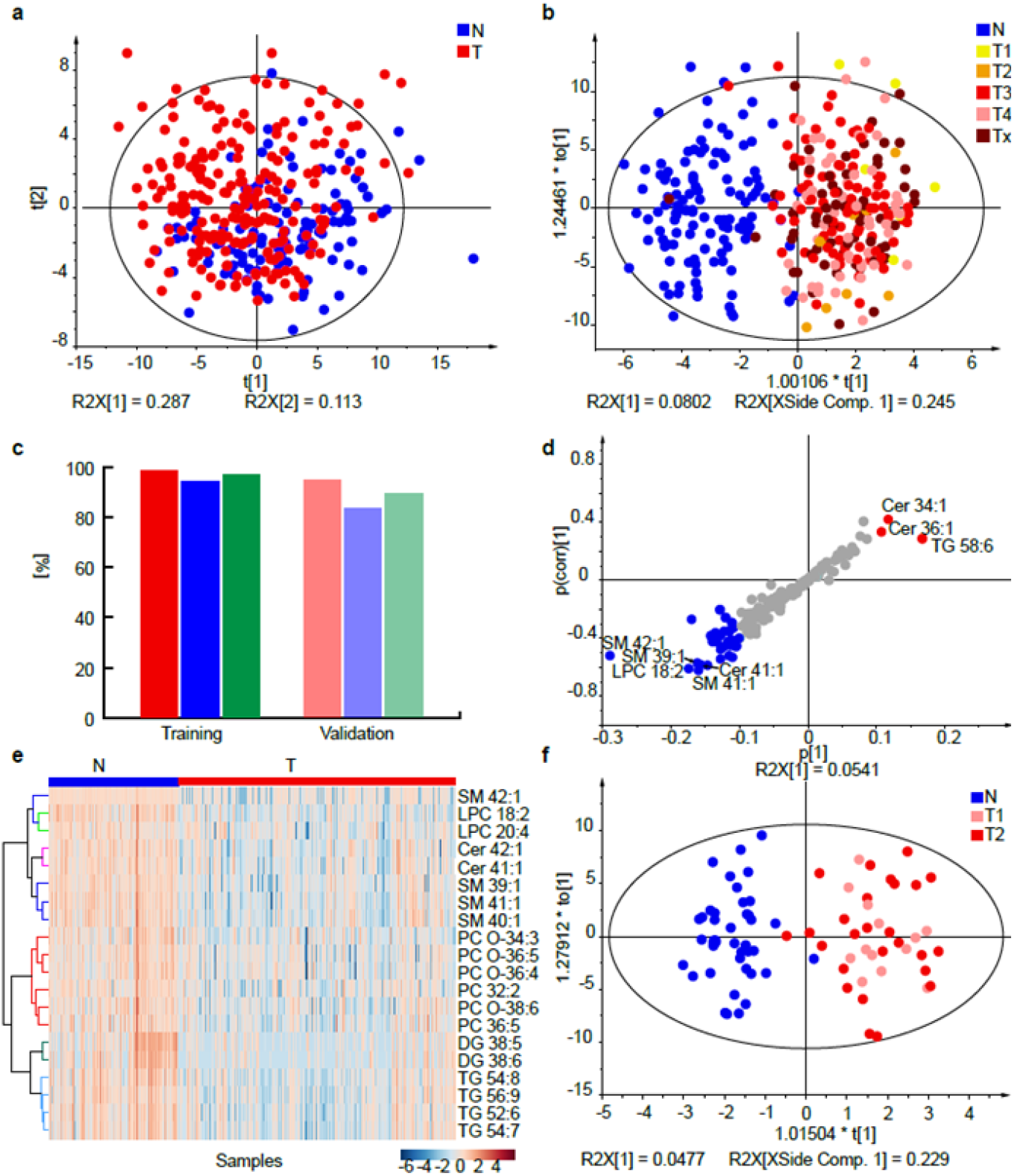
Results for the lipidomic profiling of male serum samples from PDAC patients (T) and healthy controls (N) in the Phase III. **a**, PCA for the training set (219 T + 122 N). **b**, OPLS-DA for the training set (219 T + 122 N). Individual samples are colored according to tumor (T) stage: T1 - yellow, T2 - orange, T3 - red, T4 - rose, and Tx - brown (no information about the stage was provided). **c**, Sensitivity (red), specificity (blue), and accuracy (green) for the training (219 T + 122 N) and validation (56 T + 6 N) sets. **d**, S-plot for the training set with the annotation of most up-regulated (red) and down-regulated (blue) lipid species. **e**, Heat map for both training and validation sets (275 T + 128 N). **f**, OPLS-DA for early stages T1+T2, age aligned (mean age is 65 ± 4 years for N and 67 ± 4 for T), and number aligned (39 T + 39 N). This graph includes both genders.

For PDAC screening, the key issue is the performance for early stage cancer detection, because the clinical utility of such laboratory test for late stage is not likely. Unfortunately, early stage PDAC patients typically account for a small subgroup among all cases, but all 15 males and 23 females with T1 classification in our study are correctly assigned to cancer group (Supplementary Table 10). We prepared OPLS-DA model solely for T1 and T2 tumors, merging males and females into one set to reach a sufficient number of subjects for better robustness and compared with age and number aligned healthy controls without any treatment (Fig. 3f). This model supported the observation from other graphs that early stage patients were assigned with the same accuracy as for late phases (Extended Data Fig. 4a-d and 5a-d). The prediction of health status in the validation set is based on predicted response values calculated from OPLS-DA model in SIMCA software (Supplementary Table 10), where values ≤0.5 are classified as normal, while values >0.5 are predicted as PDAC (Extended Data Fig. 7f). Regions >0.75 and <0.25 provide a very high level of confidence. On contrary, the region 0.4 – 0.6 has the higher level of uncertainty, and the majority of false classifications belongs to this middle region. The approach will be used for the future screening, when the clinician will obtain positive/negative output with based on predicted response values together with a single number from the interval <0 – 1> indicating the confidence of the prediction.

The most common chronic disease of pancreas is chronic pancreatitis, therefore concentrations of the most dysregulated lipids SM 41:1 and Cer 41:1 were compared among pancreatitis, PDAC, and healthy controls (Extended Data Fig. 8b,c). Lipid profiles of PDAC patients before and after surgery did not show any visible changes (Extended Data Fig. 8d-f), which suggests that PDAC might be a systemic disease, and that tumor removal does not cause immediate return of lipidomic profile to the premorbid condition. Medical treatment did not affect lipid profiles of serum samples either (Extended Data Fig. 8g,h). Subjects with diabetes mellitus were included in both case and control groups, and these cases do not exhibit any measurable effect on the cluster discrimination, as illustrated by box plots for SM 41:1 as the most dysregulated lipid (Extended Data Fig. 8i). OPLS-DA models (Extended Data Fig. 8j,k) were prepared for patients before any treatment and groups of age matched healthy controls to exclude any possible biases caused by treatment. The accuracy over 90% and the same patterns of dysregulated lipids show that the actual treatment did not cause relevant changes in lipid profiles.

From a biological point of view, the altered lipid metabolism may originate from tumor cells, tumor stroma, apoptotic cells, and organs affected by PDAC metastatic spread. An immune response of the organism may also be involved. All these processes can naturally contribute to the observed cancer lipidomic phenotype. In measurements from all involved laboratories, we observed a clear down-regulation of multiple lipid species in the serum of PDAC patients (Extended Data Fig. 9), such as decreased levels of most very long chain monounsaturated sphingomyelins and ceramides. These changes could be linked to the *KRAS*-driven metabolic switch^29^. In this context, alterations in sphingolipids concentrations deserve attention, as the normal metabolism of sphingomyelins might be necessary to maintain *KRAS* function^30^. Targeted biological investigations are needed to explain the mechanism of lipid alterations in the serum of PDAC patients, but it will require the development of suitable animal models in the future.

Finally, we investigated the potential of lipids for prognostic purposes using Kaplan– Meier plots, which enabled the visualization of individual parameters on the survival prognosis from the lifetime data based on non-parametric statistics. The correlation of gender (p=0.077) was not statistically significant (Extended Data Fig. 10a,b), but concentrations of the following lipids exhibited a significant effect (p<0.05) on the survival based on the data from all participating laboratories (Fig. 4 a-c, Extended Data Fig. 10b-h, and Supplementary Table 11). LPC18:2 was positively correlated with survival, which is in agreement with the previous work^13^. In contrast, Cer 36:1, Cer 38:1, Cer 42:2, PC 32:0, PC O-38:5, and SM 42:2 were negatively correlated with the survival. CA 19-9 had a strong negative correlation with the survival function (Extended Data Fig. 10i). Cox proportional-hazards model was another regression tool used for the visualization of associations among survival time and predictor variables (Fig. 4d), which demonstrated that the concentration of LPC 18:2 higher than median was positively correlated with survival, while the opposite trend was observed for CA 19-9 and PC O-38:5.

**Fig. 4.**
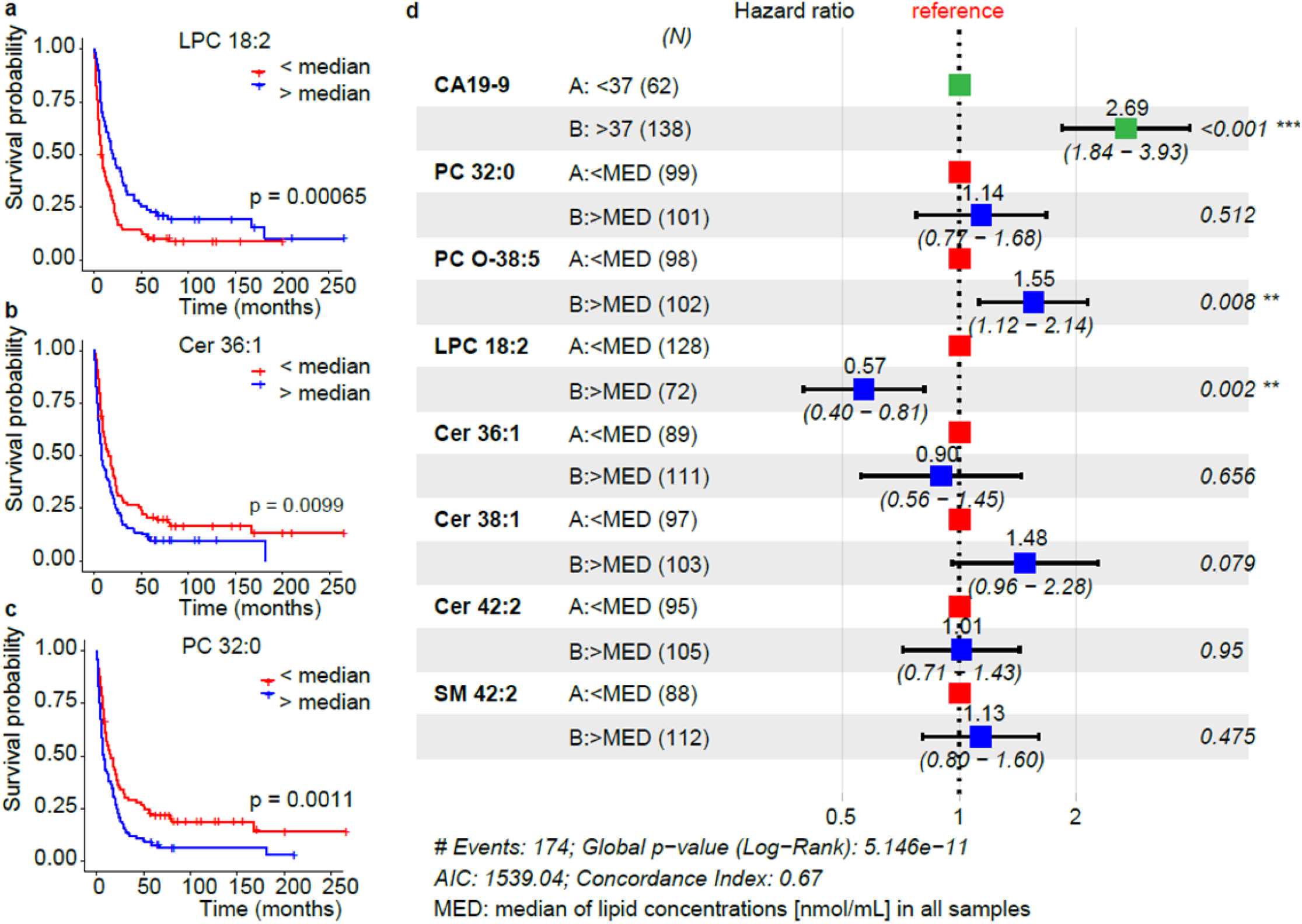
Potential of selected lipids for the survival prognosis in the Phase II measured by UHPSFC/MS. Kaplan-Meier plots for: **a**, LPC 18:2 (n=128 for binary code 0, and n=72 for binary code 1), **b**, Cer 36:1 (n=89 for 0, and n=111 for 1), and **c**, PC 32:0 (n=99 for 0, and n=101 for 1). **d**, Cox proportional-hazards model for CA 19-9, PC 32:0, PC O-38:5, LPC 18:2; Cer 36:1, Cer 38:1, Cer 42:2, and SM 42:2. Lipid species concentrations normalized to the NIST reference material obtained for all samples in the Phase II were converted into the binary code, whereby 0 was set for c < median and 1 for c > median (the median of concentrations was calculated for each lipid species including all samples).

In summary, we developed a robust and high-throughput lipidomic profiling approach for early detection of PDAC in human serum, which is applicable for screening of at least 2,000 people per month on one MS system. The real clinical utility for early PDAC screening has to be confirmed in planned large prospective cohort for high-risk individuals (hereditary PDAC, newly diagnosed diabetes mellitus in patients over 50 years and with weight loss, and patients with vague symptoms) including deeper investigation of other comorbidities. The screening lipidomic test for PDAC will have a simple readout in the form of single number, which provides a clear information on the health status for clinicians. All positive cases from the lipidomic screening test must be confirmed by conventional diagnostic approaches.

## METHODS

### Chemicals and standards

In lab 1 (University of Pardubice, Czech Republic), solvents for sample preparation and analysis, such as acetonitrile, 2-propanol, methanol (HPLC/MS grade), hexane, and chloroform stabilized with 0.5-1 % ethanol (both HPLC grade), were purchased from either Sigma-Aldrich (St. Louis, MO, USA) or Merck (Darmstadt, Germany), respectively. Mobile phase additives (ammonium acetate, ammonium formate, and acetic acid) were purchased from Sigma-Aldrich. Deionized water was obtained from a Milli-Q Reference Water Purification System (Molsheim, France). Carbon dioxide of 4.5 grade (99.995%) was purchased from Messer Group (Bad Soden, Germany). Non-endogenous lipids used as internal standards (IS) for the quantitative lipidomic analysis were purchased either from Avanti Polar Lipids (Alabaster, AL, USA), Nu-Chek (Elysian, MN, USA), or Merck. Lipid concentrations used for the IS mixture are provided in the Supplementary Table 1 depending on the employed method, further details for the preparation and dilution of the IS mixture used for UHPSFC/MS measurements were previously published^1^. The NIST SRM 1950 metabolite reference plasma was used as quality control (QC) sample and for normalization of concentrations between different MS based methods. Furthermore, a pooled serum sample of PDAC patients and healthy controls were used as QC sample. The lipid annotation used in this manuscript^2-4^ is according to the recommendations of the Lipidomics standard initiative (LSI) and given in Supplementary Table 2. The chemicals and standards mentioned above were used for the sample preparation and measurements performed in lab 1.

In lab 2 (University Hospital of Regensburg, Germany), chloroform and 2-propanol were purchased from Roth (Karlsruhe, Germany) and methanol from Merck (Darmstadt, Germany). All solvents were HPLC grade. Ammonium formate and cholesteryl ester (CE) standards were purchased from Sigma-Aldrich (Taufkirchen, Germany). TG and DG standards were purchased from Larodan (Solna, Sweden) and dissolved in 2,2,4-trimethylpenthane/2-propanol (3:1, v/v). Phosphatidylcholine (PC), ceramide (Cer), sphingomyelin (SM), lysophosphatidylcholine (LPC), and lysophosphatidylethanolamine (LPE) standards were purchased from Avanti Polar Lipids (Alabaster, Alabama, USA), and dissolved in chloroform.

In lab 3 (National University of Singapore), chemicals and reagents were obtained from the following sources: ammonium formate, acetic acid, and butanol from Sigma-Aldrich or Merck (Darmstadt, Germany); MS-grade acetonitrile and methanol from Fisher Scientific (Waltham, MA, USA); lipid standards from Avanti Polar Lipids (Alabaster, AL, USA). Ultrapure water (18 MΩ·cm at 25°C) was obtained from an Elga Labwater system (Lane End, UK).

### Phases of the study

The study is categorized into three phases in line with the recommendation in the literature^5^: Phase I (discovery), Phase II (qualification), and Phase III (verification). In Phase I, 364 samples were investigated for the lipidomic serum profile differentiation of PDAC patients from healthy controls in the main laboratory (lab 1 - Pardubice) using UHPSFC/MS. For the confirmation of results, the samples were again randomly processed and measured with shotgun MS and, for a smaller subset, with MALDI-MS in lab 1. For Phase II, new sample aliquots (554 samples) from the Masaryk Memorial Cancer Institute in Brno were obtained, further re-aliquoted, and distributed among the laboratory at University of Pardubice, Czech Republic (lab 1), laboratory at University Hospital of Regensburg, Germany (lab 2), and laboratory at National University of Singapore (lab 3). Each laboratory processed the sample set independently according to their preferred sample preparation method. For the quantitative lipidomic serum profile analysis in all three laboratories, no specifications of the applied mass spectrometry-based method were provided. The purpose was that the individual laboratories should apply their preferred, optimized, and validated methods for the lipidomic analysis. This experimental design is purposely selected to rule out that PDAC differentiation from controls and dysregulation of specific lipids is method-or laboratory-dependent. The following MS-based analytical methods were used for Phase II: UHPSFC/MS (lab 1), shotgun MS with low- and high-resolution (lab 2), and RP-UHPLC/MS (lab 3). The sample preparation protocol and lipidomic analysis was further developed and validated in the lab 1 between Phase I and Phase II, the optimized and validated conditions were applied for Phase II and III^1,6^. Phase III was performed in lab 1 using UHPSFC/MS for the serum lipidomic analysis of samples obtained from different collection sites to verify that lipidomics profiling is diagnostically conclusive and independent of the sample collection site. 554 samples from Phase II are included in 830 samples of Phase III in lab 1.

### Serum samples

Blood samples were drawn after overnight fasting. For Phase I (364 samples) and Phase II (554 samples), all human serum samples and clinical data were obtained from the Masaryk Memorial Cancer Institute in Brno, approved by the institutional ethical committees, and all blood donors signed informed consent. The sample selection was based on the availability of stored serum samples. The only exclusion criterion for healthy controls (normal, N) was the absence of malignant diseases in the life-time history without any other exclusion criteria for other diseases. For all PDAC patients, the disease was confirmed by abdominal computed tomography and/or endoscopic ultrasound followed by needle biopsy or surgical resection. All PDAC patients and healthy controls were of Caucasian ethnicity. The samples were collected from 2013 to 2015. For Phase III (830 samples), serum samples and clinical data were provided by the Masaryk Memorial Cancer Institute in Brno (554 samples, see Phase II), by the First and Third Faculty of Medicine at the Charles University in Prague (147 samples), by the University Hospital in Pilsen (31 samples) and by the Palacký University and University Hospital in Olomouc (98 samples). 22 patients with chronic pancreatitis (9 females and 13 males) treated at two outpatient departments were enrolled in this study. The etiology of pancreatitis was either ethanol-induced or recurrent acute pancreatitis. The diagnosis was confirmed by imaging methods (endoscopic ultrasound or endoscopic retrograde cholangio-pancreatography). The overview and detailed description of clinical data and patient characteristics are provided in Supplementary Tables 6 and 7. The samples were independently processed for each method used in the study. In order to avoid biases due to the sample collection, sample preparation and measurements, all samples within the particular phase were processed and measured in the randomized order. The operator had no information about the sample classification during the sample preparation and measurements. The sample sets in all phases were divided into the training and validation sets to determine the assay performance using the rigid rule defined before the study that each 6^th^ sample belongs to the validation set, and the rest constitutes the training set. The sample classification for the training set was disclosed for the multivariate data analysis (MDA). The classification of the validation set was disclosed after the final prediction of the validation set.

### Sample preparation

Briefly, the whole blood was drawn into tubes containing no anticoagulant (Sarstedt S-Monovette, Germany) and incubated at room temperature for 60 min. Then, the samples were centrifuged at 1500 × g for 15 min, the serum was isolated, immediately frozen and stored at -80°C until the extraction.

The final lipid extraction protocol in lab 1 represents a modified Folch procedure published earlier^1,7^. Human serum (25 µL) and the mixture of IS (20 µL) were homogenized in 3 mL of chloroform/methanol (2:1, v/v) for 15 min in an ultrasonic bath (40°C). When samples reached ambient temperature, 600 µL of ammonium carbonate buffer (250 mM) was added, and the mixture was ultrasonicated for 15 min. After 3 min of centrifugation (3000 rpm), the organic layer was removed, and 2 mL of chloroform was added to the aqueous phase. After 15 min of ultrasonication and 3 min of centrifugation, the organic layers were combined and evaporated under a gentle stream of nitrogen. The residue was dissolved in the mixture of 500 µL of chloroform/methanol (1:1, v/v) and vortexed. The sample preparation protocol in the Phase I was slightly different, because only single extraction was employed without any buffer, with different IS concentrations, and only vortexing instead of ultrasonication.

Finally, the extract was diluted 1:5 with chloroform/methanol (1:1, v/v) or 1:20 with the mixture of hexane/2-propanol/chloroform (7:1.5:1.5, v/v/v) (Phase I) for the UHPSFC/MS analysis, 1:8 with chloroform/methanol/2-propanol (1:2:4, v/v/v) mixture containing 7.5 mM of ammonium acetate and 1% of acetic acid for the shotgun MS analysis, and 1:1 (v/v) with methanol for the MALDI-MS.

The lipid extraction in the lab 2 was performed according to the Bligh and Dyer protocol^6^ in the presence of exogenous lipid species as IS (Supplementary Table 1c) using 10 µL of human serum for the extraction. Chloroform phase was recovered by the pipetting robot (Tecan Genesis RSP 150) and vacuum dried. Residues were dissolved in either 7.5 mM ammonium acetate in methanol/chloroform (3:1, v/v) (for low-resolution tandem mass spectrometry) or chloroform/methanol/2-propanol (1:2:4 v/v/v) with 7.5 mM ammonium formate (for high-resolution mass spectrometry).

The lipid extraction in the lab 3 was performed in a randomized order using the stratified randomization based on the sample group, age, gender, and BMI. The sample extraction was done over three days (∼230 samples / day). Human serum samples (∼100 μL each) were taken out of −80°C freezer into a biosafety cabinet and thawed on ice. 10 μL of each serum sample was transferred into 1.5 mL Eppendorf tubes. In addition, 5 μL of each serum sample was pooled together, mixed, and then 10 µL was aliquoted in 59 Eppendorf tubes to constitute batch quality control (BQC) samples. Process blanks (PBLK1-4) were prepared by aliquoting 10 µL of water into 1.5 mL Eppendorf tubes for the extraction control. 10 µL of commercial human plasma was pipetted into 1.5 mL Eppendorf tubes as reference samples (LTR1-4). 10 µL of NIST SRM 1950 plasma was pipetted into 1.5 mL Eppendorf tubes as additional reference samples (NIST1-4). The extraction was done on all above-mentioned samples as follows: Add 190 µL of chilled butanol/methanol (1:1, v/v) containing IS to the samples. Vortex each sample for 10 seconds and sonicate in ice water for 30 min. Centrifuge at 14,000 relative centrifugal force for 10 min at 4°C to pellet insoluble. Transfer 140 µL of supernatant into clean vials. Pool 30 µL of lipid extract from each vial (only from samples, not including BQC, NIST and LTR), mix, and aliquot into 59 vials as technical quality control (TQC) samples. The TQC extract was diluted with chilled butanol/methanol (1:1, v/v) to prepare 80%, 60%, 40% and 20% diluted TQC solution, which were used to assess the instrument response linearity. The lipid extracts in LC/MS vials were kept in the − 80°C freezer until LC/MS/MS analysis. On the day of analysis, LC/MS vials were taken out of the freezer, thawed at room temperature for 30 min, sonicated in ice-cold water for 15 min, and injected into LC/MS/MS.

### Measurements of CA 19-9

CA19-9, a mucin corresponding to the sialylated Lewis (Le)^a^ blood group antigen, was quantitatively determined using the electro-chemoluminescence immunoassay Elecsys^®^ (Roche, Rotkreuz, Switzerland) according to manufacturer instructions. The CA19-9 test was performed only for PDAC samples. The cut-off value for the CA 19-9 test is 37 U/mL, therefore all values over 37 U/mL were classified as PDAC.

### UHPSFC/ESI-MS conditions (lab 1)

UHPSFC/MS measurements were carried out on the Acquity Ultra Performance Convergence Chromatography (UPC^2^) system coupled to the hybrid quadrupole-traveling wave ion mobility time-of-flight mass spectrometer Synapt G2-Si from Waters by using the commercial interface kit (Waters, Milford, MA, USA). The chromatographic settings were used with minor improvements from previously published methods^1,8^. The main difference is that the data were recorded in the continuum mode. The peptide leucine enkephalin was used as the lock mass with the scan time of 0.1 s and the interval of 30 s. The lock mass was scanned but not automatically applied. The noise reduction was performed on raw files using the Waters compression tool, and then data was lock mass corrected as well as converted into the centroid data using the exact mass measure tool from Waters. For the data pre-processing, the MarkerLynx software from Waters was used. Further data processing was done by LipidQuant software available on figshare (https://figshare.com/s/cc087785ca362af7118e).

### Shotgun MS conditions (lab 1)

Shotgun experiments were performed on the quadrupole-linear ion trap mass spectrometer 6500 QTRAP (Sciex, Concord, ON, Canada) equipped with the ESI probe. Characteristic precursor ion (PI) and neutral loss (NL) scan events were used for the detection of individual lipid classes and previously reported MS settings applied^9^. For the data analysis, all observed ions in the positive-ion mode characterized by *m/z* values, type of scan, and ion intensities were exported as .txt data file and further processed using the LipidQuant software available on figshare (https://figshare.com/s/b28049603a4f361c818b).

### MALDI-MS conditions (lab 1)

MALDI matrix 9-aminoacridine (Sigma-Aldrich, St. Louis, MO, USA) was dissolved in methanol – water mixture (4:1, v/v) to the concentration of 5 mg/mL. Diluted lipid extracts of serum were mixed with matrix (1:1, v/v). The volume of 1 µL of extract/matrix mixture was deposited on the target plate using the dried droplet crystallization. A small aliquot of chloroform was applied onto MALDI plate spots before the application of diluted extract/matrix mixture to avoid the drop spreading. Mass spectra were measured on the high-resolution MALDI mass spectrometer LTQ Orbitrap XL (Thermo Fisher Scientific, Waltham, MA, USA) equipped with the nitrogen UV laser (337 nm, 60 Hz) with a beam diameter of about 80 µm × 100 µm. The LTQ Orbitrap instrument was operated in the negative-ion mode over a normal mass range *m/z* 400 - 2000 with the mass resolution 100,000 (full width at half-maximum definition at *m/z* 400). The zig-zag sample movement with 250 µm step size was used during the data acquisition. The laser energy corresponds to 15% of maximum, and 2 microscans/scan with 2 laser shots per microscan at 36 different positions were accumulated for each measurement to achieve a reproducible signal. Each sample (spotted matrix and lipid extract mixture) was spotted five times. The total acquisition time for one sample, including measurements of five consecutive spots, was 10 min. Each measurement was represented by one average MALDI-MS spectrum with thousands of *m/z* values. The automatic peak assignment was subsequently performed, and *m/z* peaks were matched with deprotonated molecules from a database created during the identification procedure using the LipidQuant software available on figshare (https://figshare.com/s/cb071be45cd91a7c90e2). This peak assignment resulted in the generation of the list of present *m/z* of studied lipids with the average intensities over all spectra, which was used for the further IS or relative normalization and the statistical evaluation.

### Shotgun MS conditions (lab 2)

The analysis of lipids was performed by the direct flow injection analysis (FIA) using a triple quadrupole (QqQ) mass spectrometer (FIA-MS/MS) and a Fourier Transform (FT) hybrid quadrupole – Orbitrap mass spectrometer (FIA-FTMS). FIA-MS/MS was performed in the positive ion mode using the analytical setup and the strategy described previously^10,11^. The fragment ion of *m/z* 184 was used for phosphatidylcholines (PC), sphingomyelins (SM)^11^, and lysophosphatidylcholines (LPC)^12^. The following neutral losses were applied for: phosphatidylethanolamines (PE) - 141, phosphatidylserines (PS) - 185, phosphatidylglycerols (PG) – 189, and phosphatidylinositols (PI) – 277 (ref.^13^). PE-based plasmalogens (PE-P) were analyzed according to the principles described by Zemski-Berry^14^. Sphingosine based ceramides (Cer) and hexosylceramides (HexCer) were analyzed using the fragment ion of *m/z* 264 (ref.^15^).

FIA-FTMS setup was described in details in previous work^16^. Triacylglycerols (TG), diacylglycerols (DG), and cholesteryl esters (CE) were recorded in the positive ion mode in *m/z* range 500 - 1000 for 1 min with a maximum injection time (IT) of 200 ms, an automated gain control (AGC) of 1·10^6^, 3 microscans, and a target resolution of 140,000 (at 200 *m/z*). The mass range of negative ion mode was split into two parts. LPC and lysophosphatidylethanolamines (LPE) were analyzed in the range *m/z* 400 - 650. PC, PE, PS, SM, and ceramides were measured in *m/z* range 520 - 960. Multiplexed acquisition (MSX) was used for [M+NH_4_]^+^ of free cholesterol (FC) (*m/z* 404.39) and D7-cholesterol (*m/z* 411.43) using 0.5 min of the acquisition time with the normalized collision energy of 10 %, IT of 100 ms, AGC of 1·10^5^, the isolation window of 1 Da, and the target resolution of 140,000. Data processing details were described in Höring *et al*. using the ALEX software^16,17^, which includes the peak assignment procedure and the intensity picking. The extracted data were exported to Microsoft Excel 2010 and further processed by the self-programmed Macros available on figshare (https://figshare.com/s/e336bdf3a52f04c2de1f).

Lipid species were annotated according to the shorthand notation of lipid structures derived from mass spectrometry^2^. For QqQ glycerophospholipid species, the annotation was based on the assumption of even numbered carbon chains only. SM species annotation is based on the assumption that a sphingoid base with two hydroxyl groups is present.

### RP-UHPLC/MS/MS conditions (lab 3)

The RP-UHPLC/MS/MS analysis was performed on the Agilent UHPLC 1290 liquid chromatography system connected to the Agilent QqQ 6495A mass spectrometer. The Agilent Eclipse Plus C18 column (100 mm × 2.1 mm, 1.8 µm) was used for the LC separation. The mobile phases A (30% acetonitrile – 20% isopropanol – 50% 10mM ammonium formate in H_2_O, v/v/v + 0.1% formic acid) and B (90% isopropanol – 9% acetonitrile – 1% 10mM ammonium formate in H_2_O, v/v/v + 0.1% formic acid) were used for both positive and negative ionization. The following gradient was applied: 0 min 15% B, 2.5 min 50% B, 2.6 min 57% B, 9 min 70% B, 9.1 min 93% B, 11 min 96% B, 11.1 min 100% B, 11.9 min 100% B, and 12.0 min 15% B, held for 3 min (total runtime of 15 min). The column temperature was maintained at 45°C. The flow rate was set to 0.4 mL/min and the sample injection volume was 2 µL.

The spray voltage was set to 3.5 kV in the positive ionization mode and 3 kV in the negative ionization mode. The nozzle voltage was set at 1 kV. The drying gas temperatures were kept at 150°C. The sheath gas temperature was 250°C. The drying gas and sheath gas flow rates were 14 L/min and 11 L/min, respectively. The nebulizer gas setting was 20 psi. The iFunnel high- and low-pressure RF were 180 V and 160 V, respectively, in the positive ionization mode and 90 V and 60 V, respectively, in the negative ionization mode. The MRM list is provided in the Supplementary Table 12.

Quantitative data were extracted by using the Agilent MassHunter Quantitative Analysis (QqQ) software. The data were manually curated to ensure that the software integrated the right peaks. Peak areas of extracted ion chromatograms peaks for each MRM transition were exported to Microsoft Excel. Peak areas were normalized to peak areas of IS using an in-house R script. The data quality was assessed using the following criteria, MRM transitions kept for the analysis had to satisfy: coefficient of variations (CoV) measured across the QC injections < 20%, linearity TQC dilution series Pearson R^2^ > 0.80, signal in processed blanks < 10% of the signal observed in the QC. Data are available at figshare: https://figshare.com/s/1fd10f273b049b93fa24

### Method validation and quality control (lab 1)

The UHPSFC/MS method was validated in line with FDA and EMA guidelines, as previously published^1^. Solvent blanks and QC samples were regularly measured after each 40 samples. For the QC samples, a pooled serum sample and the NIST SRM reference plasma sample were extracted and aliquoted. Furthermore, a mixture of naturally occurring lipid species were used as a system suitability standard. In order to assess the instrumental state, the instrument stability and the sample preparation quality, the signal response of selected endogenous lipids and the IS in all samples were monitored during the whole sequence. The signal responses of selected lipids were plotted against the number of measured samples, which allows the visualization of outliers due to the sample preparation or instrumental failures. Typically, a gradual signal drop is observed for the IS caused by contamination of the mass spectrometer over time.^6^ Furthermore, PCA for the lipidomic profiles in all samples was performed to review for outliers and the clustering of QC samples.

### Statistical analysis

SIMCA software, version 13.0 (Umetrics, Umeå, Sweden) was used to perform the unsupervised PCA with unclassified samples, and the supervised OPLS-DA with the known sample classification. Only scatter plots of the first and second components are presented in PCA score plots. OPLS-DA separates samples into known classes and can be used for the prediction. First, studied lipids were defined as variables, and samples were defined as different observations and further classified, *i*.*e*., for the health state, gender, and cancer stage. Differences in lipid profiles between genders were observed in the Phase I (Extended Data Fig. 1), therefore data sets for males and females were handled separately. The data sets were pre-treated by a logarithmic transformation, centering, scaling (unit variance (UV) or Pareto (Par) scaling), and evaluation of outliers. Logarithmic transformation was applied for each lipid species. Centering relates relative changes of a lipid species to the average, where UV or Pareto scaling compensates the concentration variance differences for lipid species. The scaling was chosen with regard to improved separation of PDAC patient and control samples and reduced number of outliers without using class information employing PCA. Pareto scaling was superior for UHPSFC/MS, MALDI-MS, low- and high-resolution shotgun MS (lab 2) and RP-UHPLC/MS (lab 3) measurements, and UV scaling for shotgun MS measurements in lab 1 during Phase I. For PCA and OPLS-DA, the number of components was assessed by model fit and prediction ability. In the case of too few components, the differentiation of classes (*i*.*e*., health state) is insufficient, while in the case of too many components, the model may be overfitted, resulting in diminished prediction power. The model fit was determined by the evaluation of R2, which describes the variation of variables (lipid species) explained by the model. The prediction ability of the model is described by Q2 and is estimated using a cross-validation. Cross-validation was performed by dividing the data set into 7 groups, omitting one group, building the model, and predicting the omitted group. This was repeated for each group, and the results of the prediction were summarized by the variable Q2. For building models, components were added as long as Q2 was increasing with the number of components. Finally, PCA plot was evaluated for outliers, errors in measurements, clustering of QC samples as well as for the separation of sample types, *i*.*e*., PDAC patients vs. healthy controls. Afterwards, OPLS-DA was performed in order to discriminate between PDAC patients and healthy controls. The number of predictive and orthogonal components for all methods is provided in Supplementary Table 8b. A confidence level on parameters of 95% was used for all models.

OPLS models were built for the training set for individual methods and validated by the prediction of the validation set using predicted response values. The unpredicted original value of Y is 0, if a human subject is without cancer, and 1 in case of PDAC (binary variable). Predicted response value is continuous and computed using the last model component. Based on the predicted value of Y, the sample is classified as non-cancerous subject (if predicted Y ≤0.5) or cancerous subject (if predicted Y>0.5). A summary of predicted response values obtained for the training and validation sets with the various methods at the different clinical phases is provided in the Supplementary Table 10. Depending on the correctly identified healthy and cancerous samples, the selectivity, specificity, and accuracy of the model for the training and validation samples were determined (Supplementary Table 8a).

In order to evaluate lipids of statistical significance, a two-sided two sample T-test assuming unequal variances (Welch test) was performed for healthy and cancerous samples. P-values <0.05 were considered to indicate the statistical significance. The Bonferroni approach was applied to all p-values for the multiple testing correction. The summary statistics and average molar lipid concentrations for healthy and cancerous samples are summarized in the Supplementary Tables 9a-c for all methods and study phases. Furthermore, the parameter of variable influence of projection (VIP) was evaluated for each statistical OPLS-DA model using the SIMCA software. Finally, only lipid species with p-values <0.05, VIP values >1, and fold changes ≥20% for molar concentrations were considered as statistically important and reported in Supplementary Tables 9a-c. For the visualization of differences in lipid concentrations (up- and down-regulation) between cancer and control samples, box plots were constructed in R free software environment (https://www.r-project.org) using ggplot2, ggpubr, and rstatix packages. In each boxplot, the median was presented by a horizontal line, box represented 1^st^ and 3^rd^ quartile values, and whiskers stood for 1.5*IQR from the median. Each measurement was plotted as jittered point value. The receiver operating characteristics (ROC) curves were generated by using the package AUC in R.

For verification of the data processing, statistical analysis and results, data were cross-checked and independently reprocessed or evaluated by applying the online metabolomics platform MetaboAnalyst (ver. 4.0)^18^.

For the Kaplan-Meier survival analysis and the Cox Hazard proportional analysis, lipid concentrations were converted into the binary code. Therefore, the median concentration of the lipid species for all samples were calculated, and individual lipid species concentrations were classified to 0, when the concentration was smaller than the median concentration of all samples or 1, when the concentration was bigger than the median concentration of all samples. The Kaplan-Meier survival analysis plots and the Cox-Hazard proportional analysis plots were generated by using the packages survival and survminer in R software.

### Outlier inspection

The QC system and the PCA analysis revealed outliers. In Phase I, sample No. 355 was excluded from the UHPSFC/MS data set, and sample No. 210 for the shotgun MS data set, due to the sample preparation failure. The repetition of the sample preparation was not possible due to insufficient serum volume. In the Phase II, samples No. 246 and 500 were excluded from the low resolution shotgun MS data set, and samples No. 246 and 409 from the high resolution shotgun MS data set.

### Data availability

All data necessary to support the conclusions are available in the manuscript or supplementary information. Raw data, instructions for the software handling and the software are deposited at figshare.com:

https://figshare.com/s/5ddbcbf1be4a1aec966f

https://figshare.com/s/b28049603a4f361c818b

https://figshare.com/s/40f1450376cdc8d69e9a

https://figshare.com/s/cb071be45cd91a7c90e2

https://figshare.com/s/1fd10f273b049b93fa24

https://figshare.com/s/e336bdf3a52f04c2de1f

https://figshare.com/s/cc087785ca362af7118e

## Supporting information

Supplementary tables guide

## Data Availability

All data necessary to support the conclusions are available in the manuscript or supplementary information. Raw data, instructions for the software handling and the software are deposited at figshare.com.

https://figshare.com/s/5ddbcbf1be4a1aec966f

https://figshare.com/s/b28049603a4f361c818b

https://figshare.com/s/40f1450376cdc8d69e9a

https://figshare.com/s/cb071be45cd91a7c90e2

https://figshare.com/s/1fd10f273b049b93fa24

https://figshare.com/s/e336bdf3a52f04c2de1f

https://figshare.com/s/cc087785ca362af7118e

## Acknowledgments

The project was funded by the project 18-12204S (the Czech Science Foundation). L.K. acknowledges the support of institutional program of the Charles University in Prague (UNCE 204064). R.H. acknowledges the support of MEYS – BBMRI-CZ LM2018125 (the Ministry of Education, Youth and Sports of the Czech Republic) and MH CZ - DRO (MMCI, 00209805, the Ministry of Health of the Czech Republic) projects.

## Author contributions

M.H., D.W., R.J., and E.C. prepared the design of experiments. D.W., M. Ch., and O.P. performed sample preparation. D.W. analyzed sample by UHPSFC/MS, E.C. by shotgun MS, and R.J. by MALDI-MS. M. Hör., D.M., G.L., R.B., M.R.W., and A.C.G. performed analysis and data evaluation in cooperating laboratories. E.C. developed the software for data evaluation. D.W., R.J., T.H., E.C., J.I., and G.V.T. processed data and performed statistical analysis. L.K., R.K., D.F., and R.A. discussed observed changes. R.H., P.K., I.N., P.Š., J.Š., R.K., and B.M. obtained and provided serum samples and corresponding data. M.H., D.W., and R.J. prepared the first draft of manuscript. D.W., R.J., J.I., and R.K. prepared figures. M.H. was responsible for funding and supervision of this study. All co-authors read and approved the manuscript.

## Extended Data - Figures legends

**Extended Data Fig. 1.**
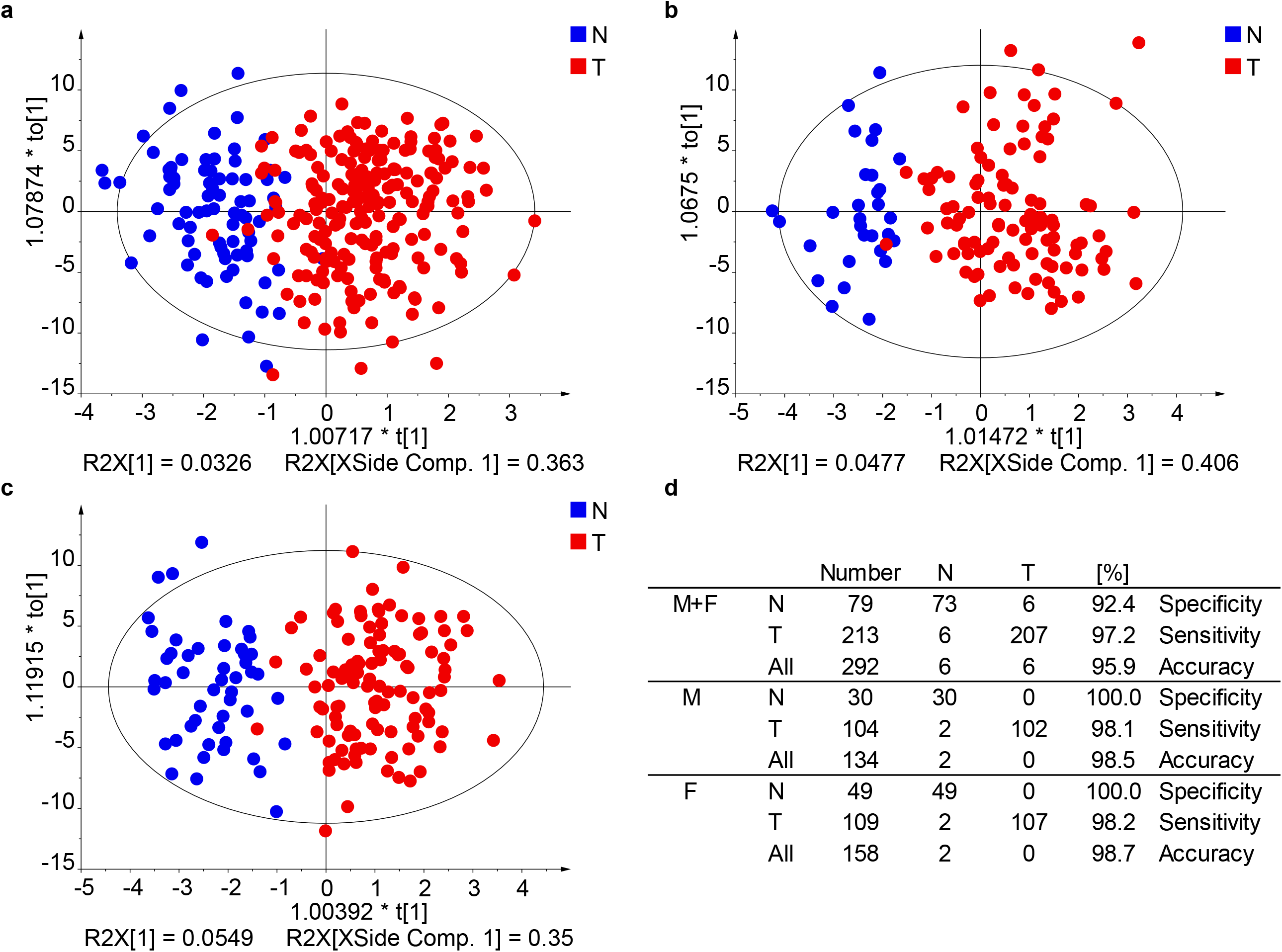
Effect of gender separation on the quality of OPLS-DA models used for the differentiation of human serum samples obtained from PDAC patients (T) and healthy controls (N) for the training set using UHPSFC/MS in the Phase I. **a**, Both genders. **b**, Males. **c**, Females. **d**, Specificity, sensitivity, and accuracy for individual models.

**Extended Data Fig. 2.**
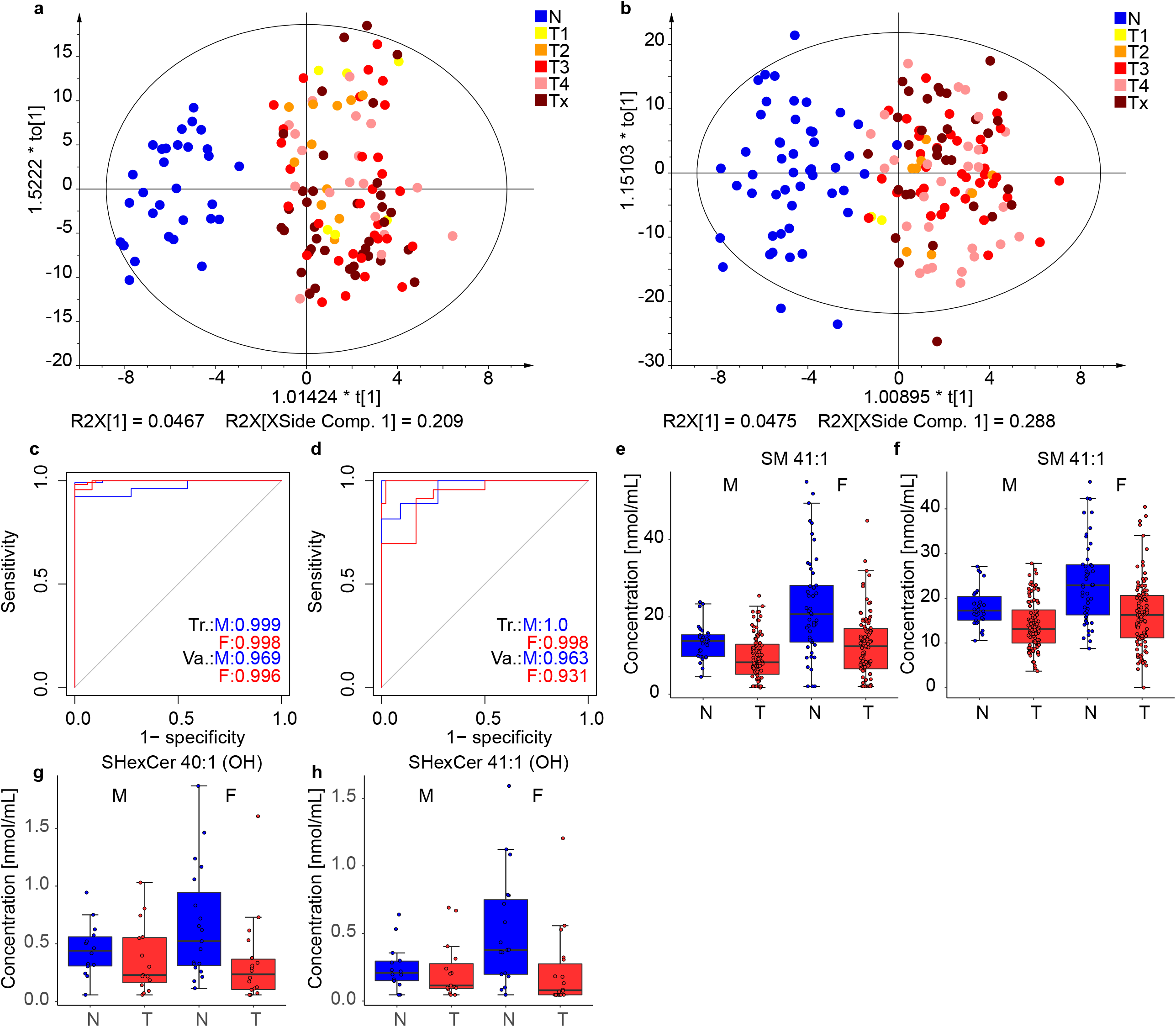
Results for the Phase I obtained in the lab 1. Individual samples are colored according to tumor (T) stage: T1 - yellow, T2 - orange, T3 - red, T4 - rose, and Tx - brown (no information about the stage was provided). **a**, OPLS-DA for males measured with shotgun MS for the training set (104 T + 30 N). **b**, OPLS-DA for females measured with shotgun MS for the training set (157 T + 49 N). ROC curves for males (M) and females (F) in training (Tr.) and validation (Va.) sets: **c**, UHPSFC/MS, and **d**, shotgun MS. Box plots for molar concentration in human serum from PDAC patients (T) and healthy controls (N) for males (M) and females (F): **e**, SM 41:1 measured by UHPSFC/MS, **f**, SM 41:1 measured by shotgun MS (LR), for both box plots for males (104 T and 30 N) and females (109 T and 49 N), **g**, SHexCer 41:1(OH) measured by MALDI-MS, and **h**, SHexCer 40:1(OH) measured by MALDI-MS, for both box plots for males (15 T and 14 N) and females (18 T and 19 N)

**Extended Data Fig. 3.**
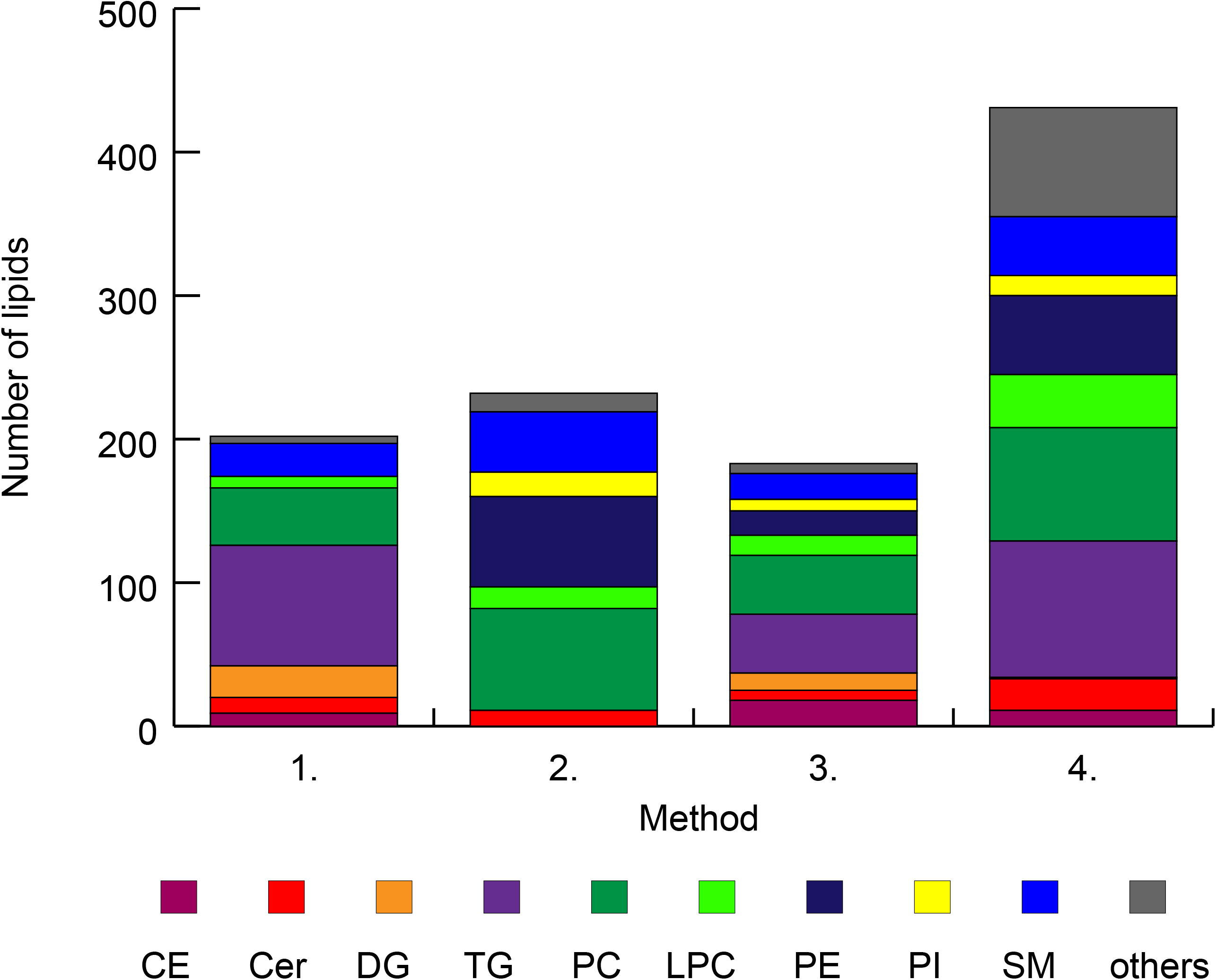
Summary of quantified lipid species for particular lipid classes. Method 1 – UHPSFC/MS measured by lab 1 (n=202), Method 2 – shotgun MS with low-resolution (LR) measured by lab 2 (n=232), Method 3 – shotgun MS with high-resolution (HR) measured by lab 2 (n=183), and Method 4 – RP-UHPLC/MS measured by lab 3 (n=431). Annotation of lipid classes: CE – cholesteryl esters, Cer – ceramides, DG – diacylglycerols, TG – triacylglycerols, PC – phosphatidylcholines, LPC – lysophosphatidylcholines, PE – phosphatidylethanolamines, PI – phosphatidylinositols, and SM – sphingomyelins.

**Extended Data Fig. 4.**
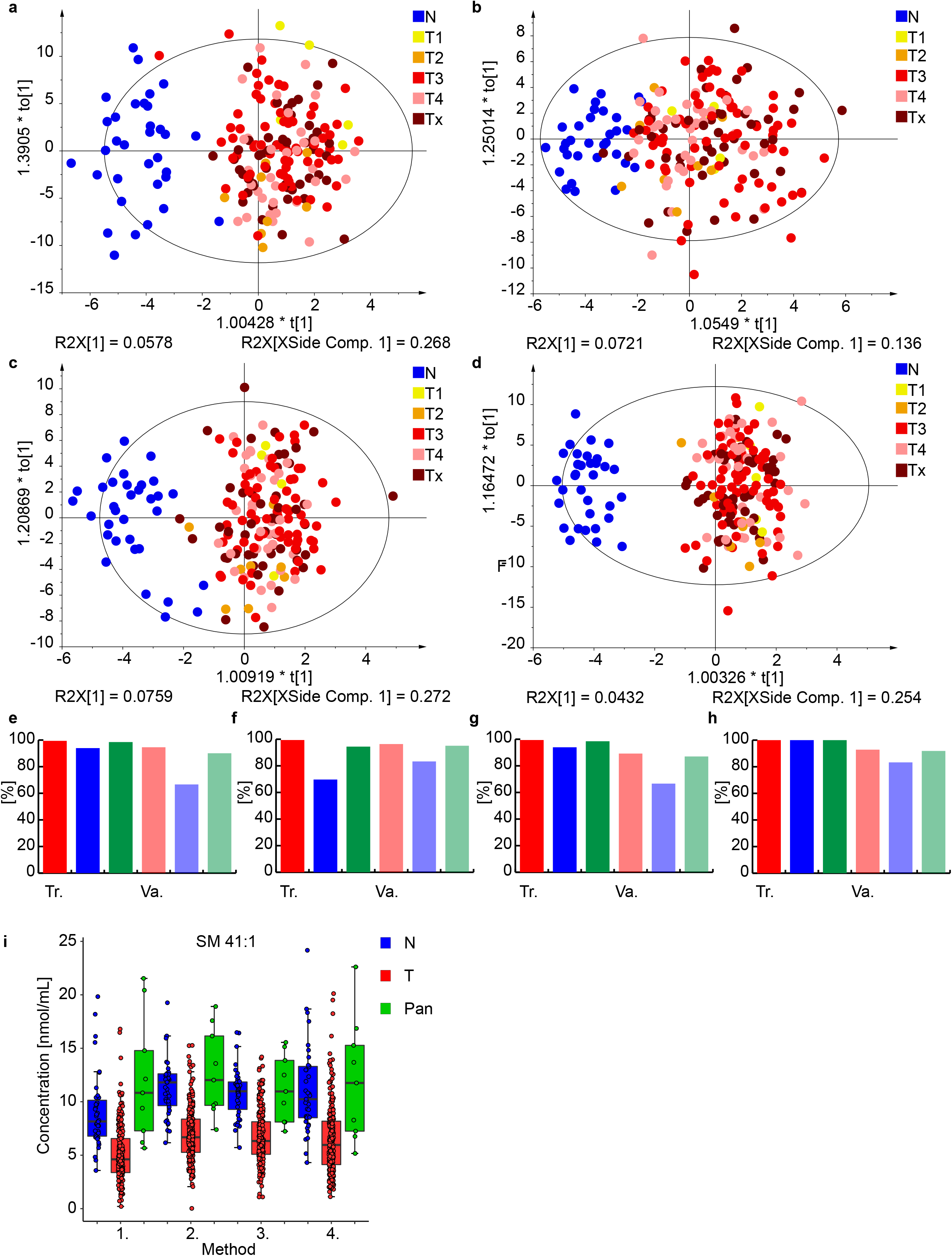
Results for the lipidomic profiling of male serum samples from PDAC patients and healthy controls in the Phase II. OPLS-DA for molar concentrations of lipid species obtained for the training set: **a**, UHPSFC/MS (166 T + 33 N), **b**, shotgun MS (LR) (165 T + 33 N), **c**, shotgun MS (HR) (164 T + 33 N), and **d**, RP-UHPLC/MS (166 T + 33 N). Individual samples are colored according to their tumor (T) stage: T1 – yellow, T2 – orange, T3 – red, T4 – rose, Tx – brown (no information about the stage was provided). Sensitivity (red), specificity (blue), and accuracy (green) values in percentage for the training (Tr.) and validation (Va.) sets: **e**, UHPSFC/MS, **f**, shotgun MS (LR), **g**, shotgun MS (HR), and **h**, RP-UHPLC/MS. **i**, Box plots of molar lipid concentrations normalized with the NIST reference material determined in PDAC patients (222 T), controls (39 N), and pancreatitis (9 Pan) patients including both validation and training sets for SM 41:1 using UHPSFC/MS (Method 1), shotgun MS (LR) (Method 2), shotgun MS (HR) (Method 3), and RP-UHPLC/MS (Method 4).

**Extended Data Fig. 5.**
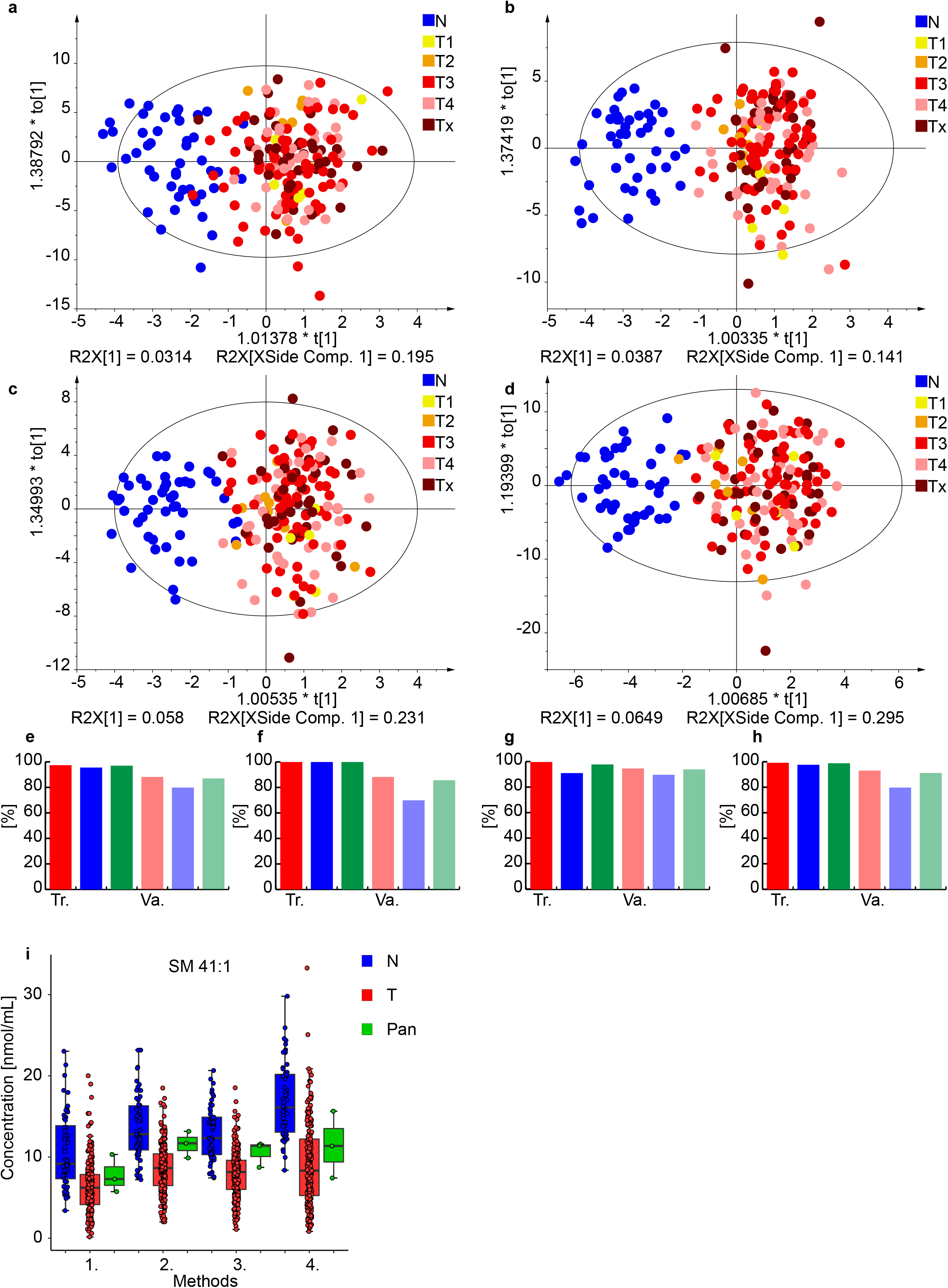
Results for the lipidomic profiling of female serum samples from PDAC patients (T) and healthy controls (N) in the Phase II. OPLS-DA for molar concentrations of lipid species obtained for the training set: **a**, UHPSFC/MS (161 T + 46 N), **b**, shotgun MS (LR) (160 T + 46 N), **c**, shotgun MS (HR) (161 T + 46 N), and **d**, RP-UHPLC/MS (161 T + 46 N). Individual samples are colored according to their tumor (T) stage: T1 - yellow, T2 - orange, T3 - red, T4 - rose, and Tx - brown (no information about the stage was provided). Sensitivity (red), specificity (blue), and accuracy (green) values in percentage for the training and validation sets: **e**, UHPSFC/MS, **f**, shotgun MS (LR), **g**, shotgun MS (HR), and **h**, RP-UHPLC/MS. **i**, Box plots of molar lipid concentrations normalized with the NIST reference material determined in PDAC patients (221 T), controls (56 N), and pancreatitis (3 Pan) patients including both validation and training sets for SM 41:1 using UHPSFC/MS (Method 1), shotgun MS (LR) (Method 2), shotgun MS (HR) (Method 3), and RP-UHPLC/MS (Method 4).

**Extended Data Fig. 6.**
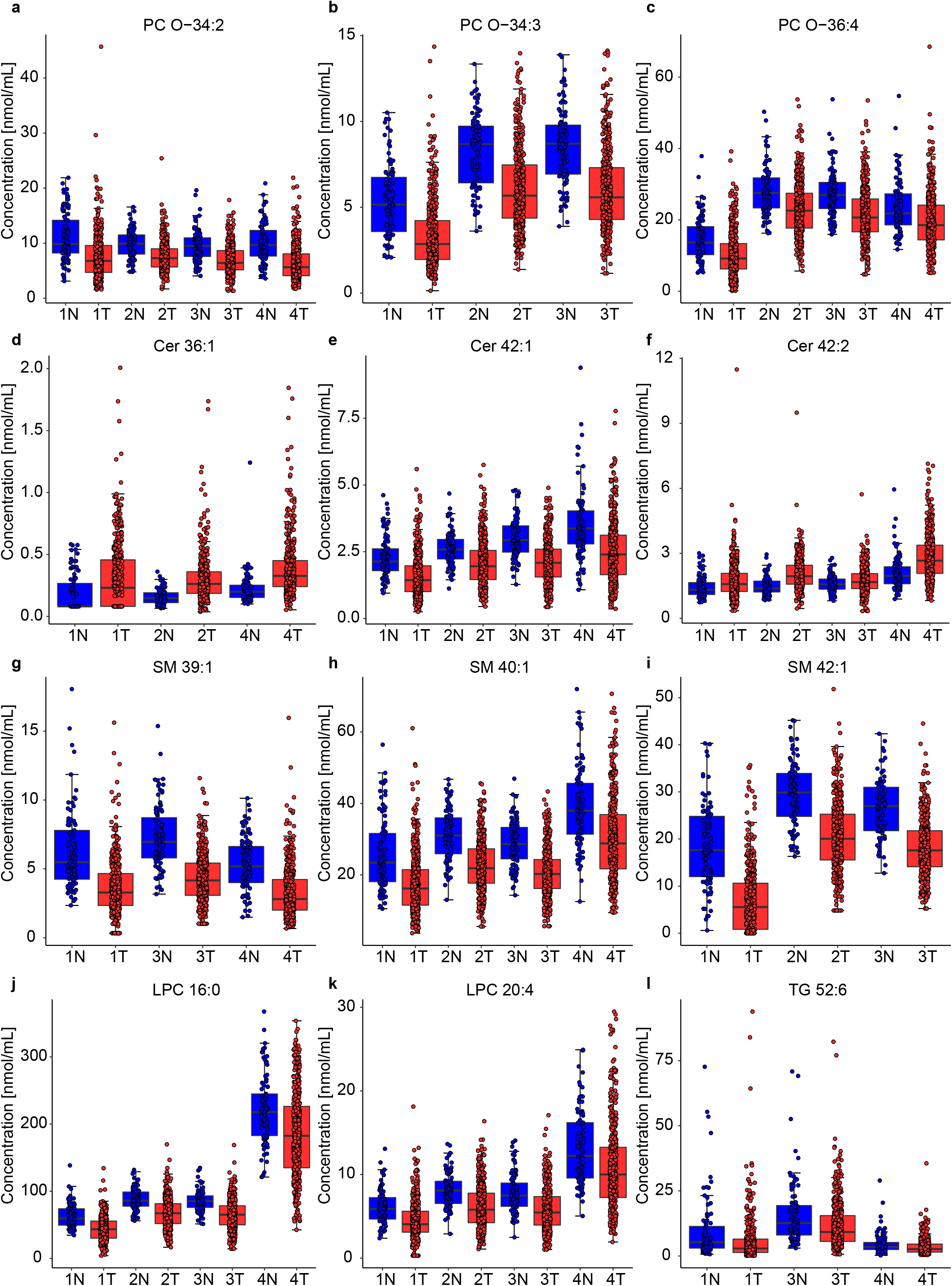
Selected box plots for the Phase II. Lipid concentrations normalized with the NIST reference material determined in PDAC patients (443 T) and healthy controls (95 N) including both validation and training sets and both genders: **a**, PC O-34:2, **b**, PC O-34:3, **c**, PC O-36:4, **d**, Cer 36:1, **e**, Cer 42:1, **f**, Cer 42:2, **g**, SM 39:1, **h**, SM 40:1, **i**, SM 42:1, **j**, LPC 16:0, **k**, LPC 20:4, and **l**, TG 52:6 for UHPSFC/MS (Method 1), shotgun MS (LR) (Method 2), shotgun MS (HR) (Method 3), and RP-UHPLC/MS (Method 4).

**Extended Data Fig. 7.**
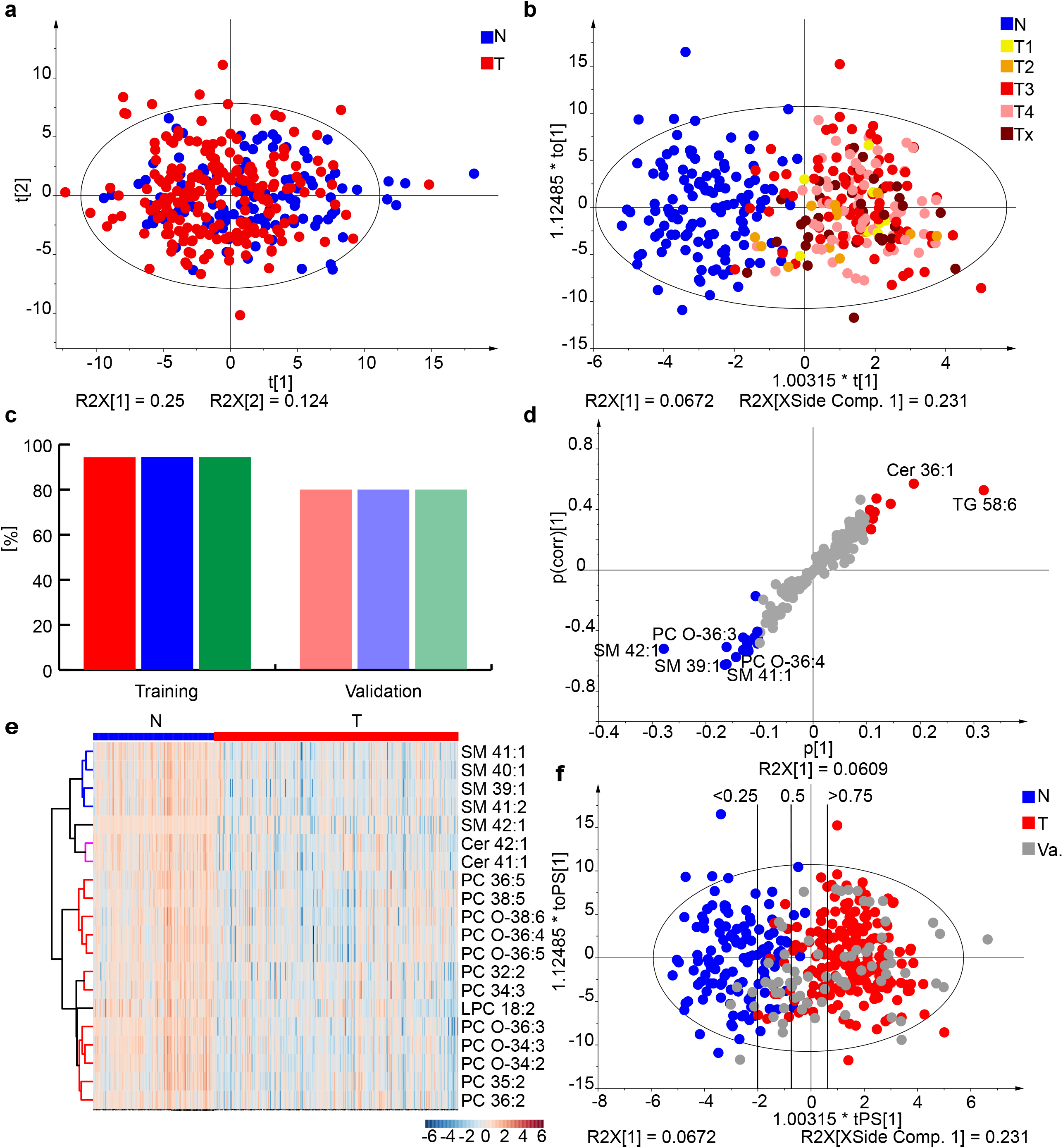
Results for the lipidomic profiling of female serum samples from PDAC patients (T) and healthy controls (N) in the Phase III. **a**, PCA for the training set (211 T + 124 N). **b**, OPLS-DA for the training set (211 T + 124 N). Individual samples are colored according to their tumor (T) stage: T1 - yellow, T2 - orange, T3 - red, T4 - rose, and Tx - brown (no information about the stage was provided). **c**, Sensitivity (red), specificity (blue), and accuracy (green) for the training and validation sets. **d**, S-plot for the training set with the annotation of most up-regulated (red) and down-regulated (blue) lipid species. **e**, Heat map for both training and validations sets (271 T + 134 N).

**Extended Data Fig. 8.**
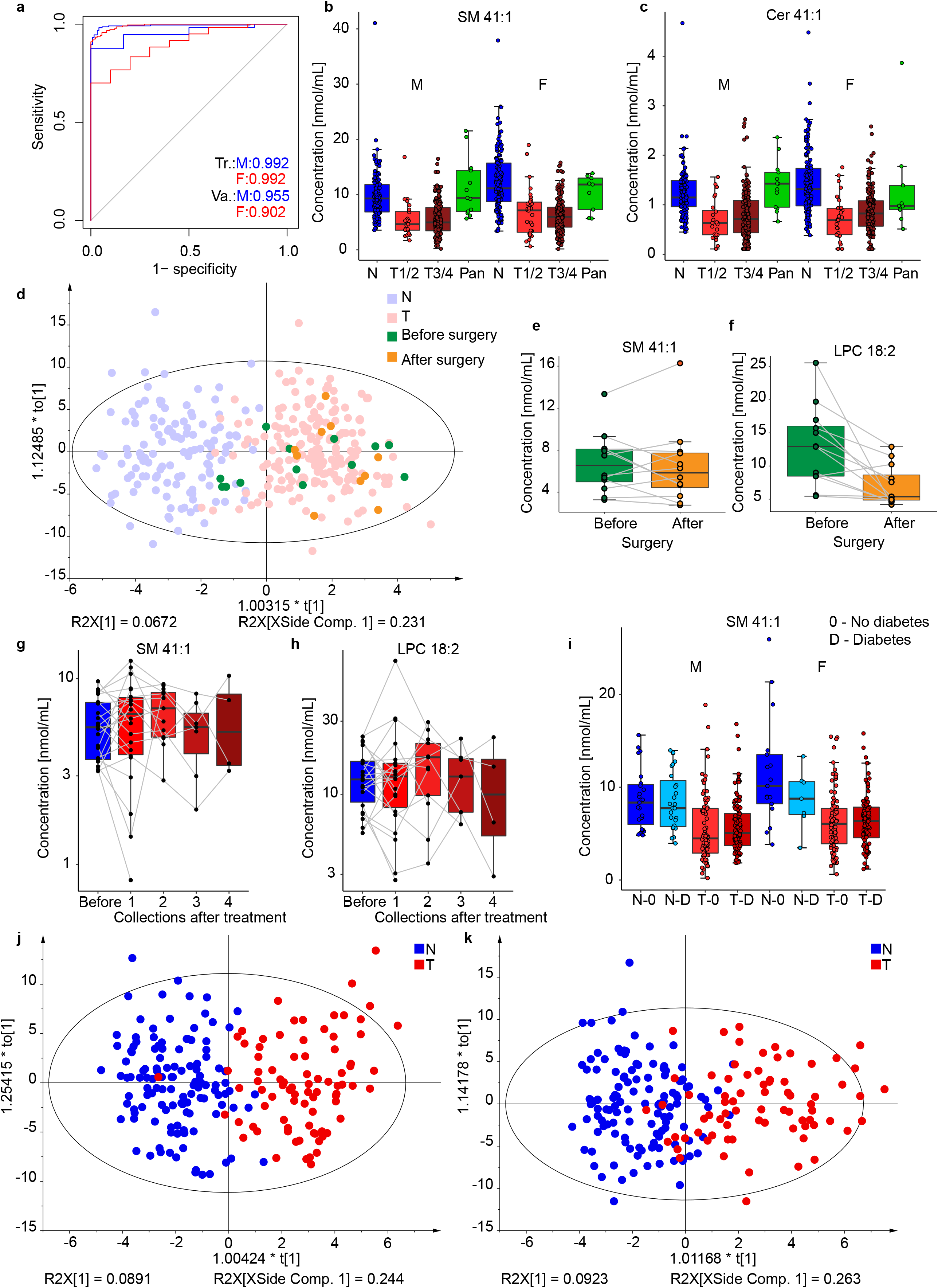
Results for the lipidomic profiling of human serum samples for PDAC patients (T) and healthy controls (N) including both genders in the Phase III. **a**, ROC curves for males (M) and females (F) in training (Tr.) and validation (Va.) sets. Box plots of lipid molar concentrations normalized using the NIST reference material for: **b**, SM 41:1, and **c**, Cer 41:1. Only samples with known tumor (T) stage classification were included, where early stages (T1 and T2, 24 males and 30 females) and late stages (T3 and T4, 174 males and 176 females) are summarized and compared to samples of healthy controls (128 males and 134 females) and pancreatitis patients (13 males and 9 females). Influence of surgery on the lipidomic profile: **d**, OPLS-DA for females (211 T + 124 N) using the training set with highlighted samples before (green, n=13) and after (orange, n=10) surgery. Box plots of molar lipid concentrations for paired samples collected before and after surgery for both genders (2 males and 10 females): **e**, SM 41:1, and **f**, LPC 18:2. Box plots for paired samples collected before (n=22) and after treatment (n=22 for collection 1, n=12 for collection 2, n=7 for collection 3, n=4 for collection 4) for both genders using molar concentrations: **g**, SM 41:1, **h**, LPC 18:2, and **i**, Cer 41:1. OPLS-DA modes only for subjects before any treatment or surgery separately for **j**, males (83 T + 122 N) and **k**, females (73 T + 124 N).

**Extended Data Fig. 9.**
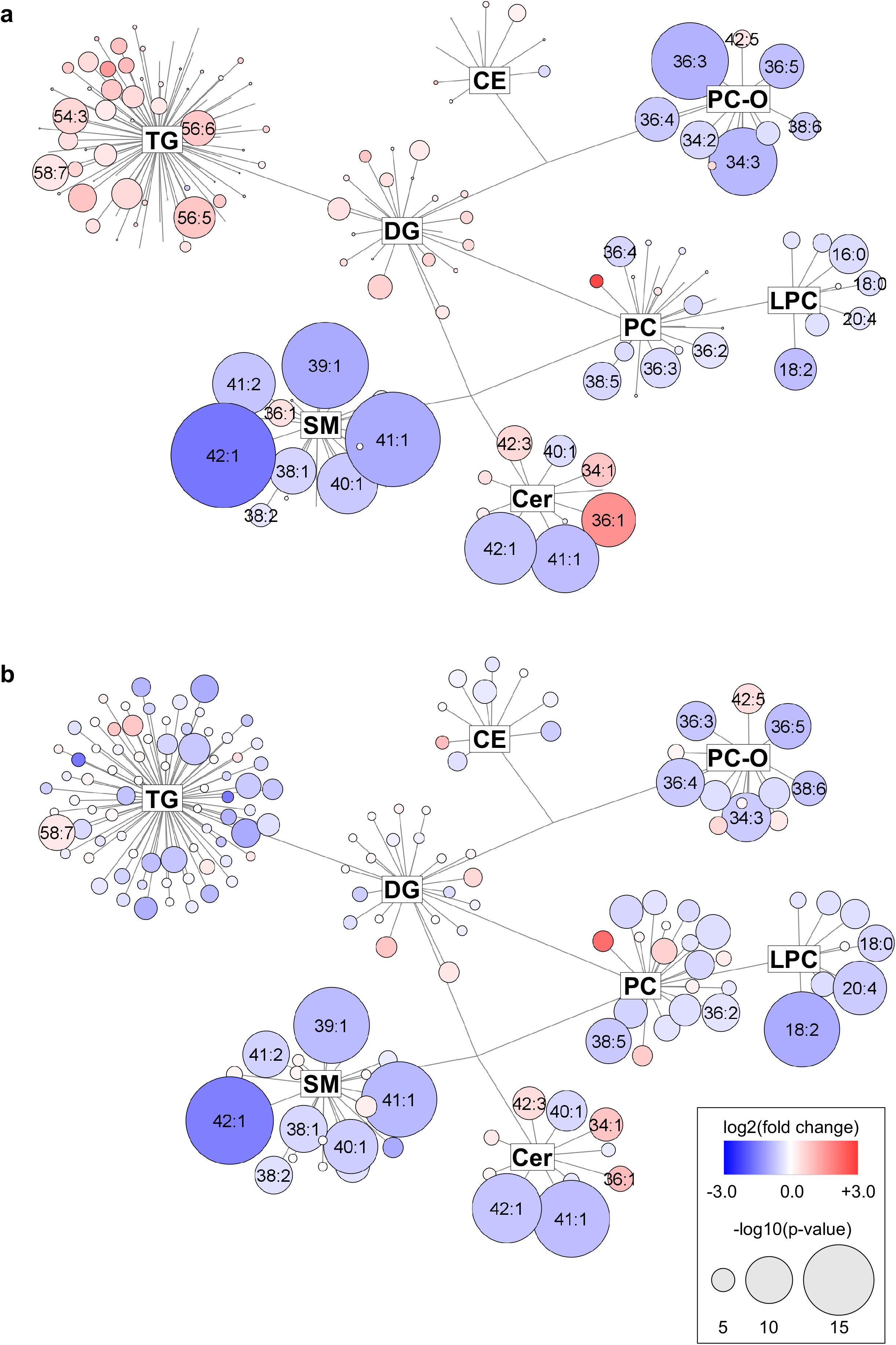
Network visualization of the most dysregulated lipid species in PDAC for data from Phase III. Graphs show lipidomic pathways with clustering into individual lipid classes for **a**, males, and **b**, females using the Cytoscape software (http://www.cytoscape.org). Circles represent detected lipid species, where the circle size expresses the significance according to p-value, while the color darkness defines the degree of up-/down-regulation (red/blue) according to the fold change. The most discriminating lipids are annotated.

**Extended Data Fig. 10.**
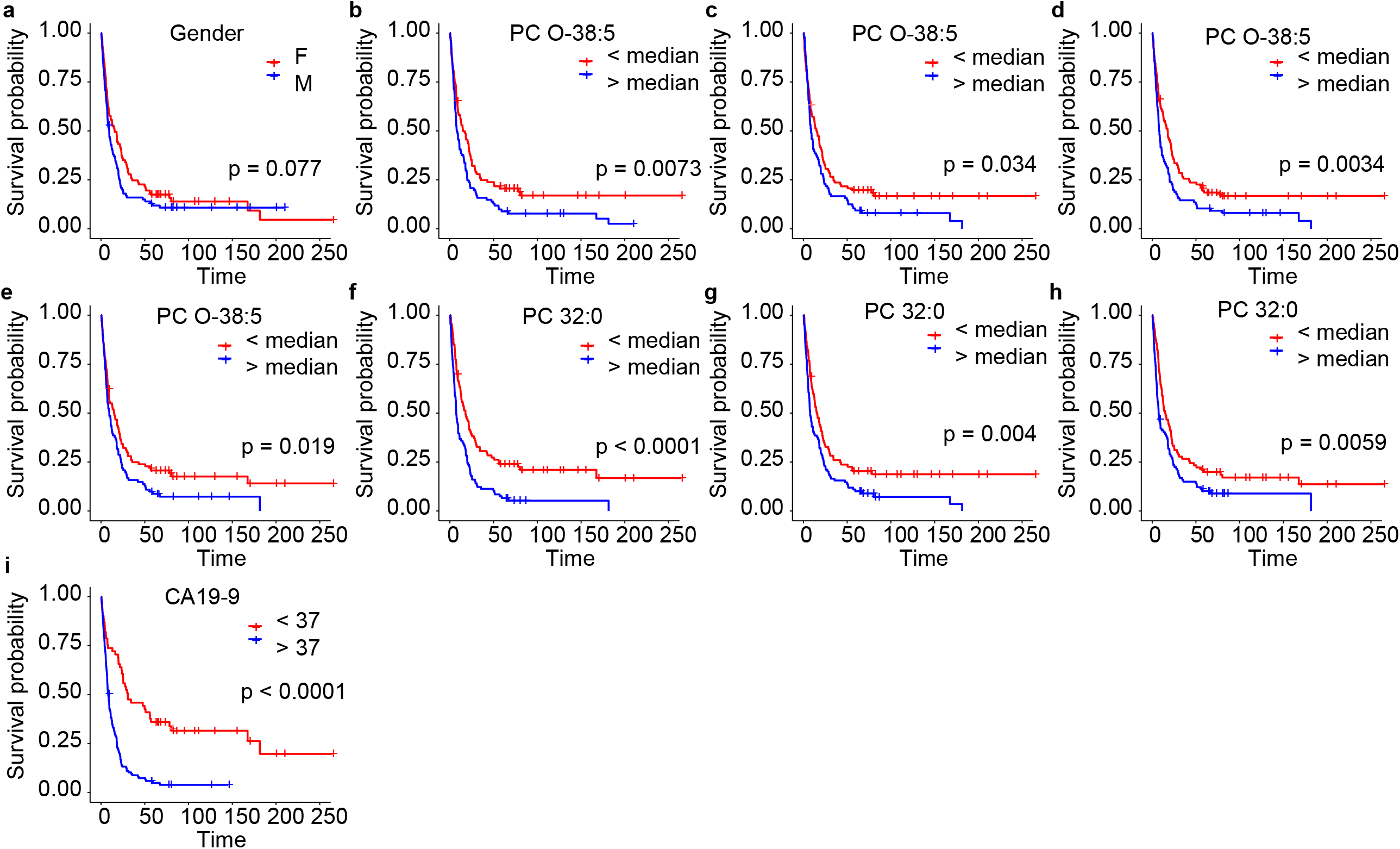
Potential of selected dysregulated lipid species for the survival prognosis in the Phase II using Kaplan-Meier plots. **a**, Gender (102 males and 98 females). **b**, PC O-38:5 measured by UHPSFC/MS (n=98 for binary code 0, and n=102 for binary code 1). **c**, PC O-38:5 measured by shotgun MS (LR) (n=103 for 0, and n=97 for 1). **d**, PC O-38:5 measured by shotgun MS (HR) (n=103 for 0, and n=97 for 1). **e**, PC O-38:5 measured by RP-UHPLC/MS (n=98 for 0, and n=102 for 1). **f**, PC 32:0 measured by shotgun MS (LR) (n=94 for 0, and n=106 for 1). **g**, PC 32:0 measured by shotgun MS (HR) (n=91 for 0, and n=109 for 1). **h**, PC 32:0 measured by RP-UHPLC/MS (n=90 for 0, and n=110 for 1). **i**, CA 19-9 (n=62 for 0, and n=138 for 1).

## References

1. Hur, C. et al. Early pancreatic ductal adenocarcinoma survival is dependent on size positive implications for future targeted screening. Pancreas 45, 1062–1066 (2016).

2. Ryan, D.P., Hong, T.S. & Bardeesy, N. Pancreatic adenocarcinoma. N. Engl. J. Med. 371, 1039–1049 (2014).

3. Wolrab, D. et al. Oncolipidomics: Mass Spectrometric Quantitation of Lipids in Cancer Research. Trends Anal. Chem. 120, 115480 (2019).

4. Holčapek, M., Liebisch & G., Ekroos, K. Lipidomic analysis. Anal. Chem. 90, 4249– 4257 (2018).

5. Cancer treatment & survivorship facts & figures. American cancer society. https://www.cancer.org/research/cancer-facts-statistics/survivor-facts-figures.html (2016-2017).

6. Toft, J. et al. Imaging modalities in the diagnosis of pancreatic adenocarcinoma: A systematic review and meta-analysis of sensitivity, specificity and diagnostic accuracy. Eur. J. Radiol. 92, 17–23 (2017).

7. Duffy, M.J. et al. Tumor markers in pancreatic cancer: a European group on tumor markers (EGTM) status report. Ann. Oncol. 21, 441–447 (2010).

8. Root, A., Allen, P., Tempst, P. & Yu, K. Protein biomarkers for early detection of pancreatic ductal adenocarcinoma: progress and challenges. Cancers 10, 67–78 (2018).

9. Cohen, J.D. et al. Detection and localization of surgically resectable cancers with a multi-analyte blood test. Science 359, 926–930 (2018).

10. Buscail, L., Bournet, B. & Cordelier, P. Role of oncogenic KRAS in the diagnosis, prognosis and treatment of pancreatic cancer. Nat. Rev. Gastroenterol. Hepatol. 17, 153–168 (2020).

11. Jones S., et al. Core signaling pathways in human pancreatic cancers revealed by global genomic analyses. Science 321, 1801–1806 (2008).

12. Bryant, K., Mancias, J., Kimmelman, A. & Der, C. KRAS: feeding pancreatic cancer proliferation. Trends Biochem. Sci. 39, 91–100 (2014).

13. Kamphorst, J. et al. Hypoxic and Ras-transformed cells support growth by scavenging unsaturated fatty acids from lysophospholipids. Proc. Natl. Acad. Sci. USA. 110, 8882–8887 (2013).

14. Salloum, D. et al. Mutant ras elevates dependence on serum lipids and creates a synthetic lethality for rapamycin. Mol. Cancer Ther. 13, 733–741 (2014).

15. Rozeveld, C.N., Johnson, K.M., Zhang, L., Razidlo, G.L. KRAS Controls Pancreatic Cancer Cell Lipid Metabolism and Invasive Potential through the Lipase HSL. Cancer Res. 80, 4932–4945 (2020).

16. Wenk, M.R. Lipidomics: new tools and applications. Cell 143, 888–895 (2010).

17. Cífková, E. et al. Correlation of lipidomic composition of cell lines and tissues of breast cancer patients using hydrophilic interaction liquid chromatography – electrospray ionization mass spectrometry and multivariate data analysis. Rapid Commun. Mass Spectrom. 31, 253–263 (2017).

18. Cífková, E. et al. Determination of lipidomic differences between human breast cancer and surrounding normal tissues using HILIC-HPLC/ESI-MS and multivariate data analysis. Anal. Bioanal. Chem. 407, 991–1002 (2015).

19. Bandu, R., Mok, H.J. & Kim, K.P. Phospholipids as cancer biomarkers: mass spectrometry-based analysis. Mass Spectrom. Rev. 37, 107–138 (2018).

20. Heiskanen, L.A., Suoniemi, M., Ta, H.X., Tarasov, K. & Ekroos, K. Long-term performance and stability of molecular shotgun lipidomic analysis of human plasma samples. Anal. Chem. 85, 8757–8763 (2013).

21. Rifai, N., Gillette, M.A., Carr, S.A. Protein biomarker discovery and validation: the long and uncertain path to clinical utility. Nat. Biotechnol. 24, 971–983 (2006).

22. Guideline on bioanalytical method validation. Committee for medicinal products for human use (CHMP). http://www.ema.europa.eu/ema/index.jsp?curl=pages/includes/document/document_detail.jsp?webContentId=WC500109686%26mid=WC0b01ac058009a3dc (First published 2011, last updated 2015).

23. Food and drug administration guidance for industry: bioanalytical method validation. US Department of health and human services. Center for drug evaluation and research: Rockville, MD. https://www.fda.gov/ForIndustry/IndustryNoticesandGuidanceDocuments/default.htm (2001).

24. Wolrab, D. et al., Determination of one year stability of lipid plasma profile and comparison of blood collection tubes using UHPSFC/MS and HILIC-UHPLC/MS. Anal. Chim. Acta 1137, 74–84 (2020).

25. Lísa, M. & Holčapek, M. High-throughput and comprehensive lipidomic analysis using ultrahigh-performance supercritical fluid chromatography–mass spectrometry. Anal. Chem. 87, 7187–7195 (2015).

26. Jirásko, R. et al. MALDI Orbitrap mass spectrometry profiling of dysregulated sulfoglycosphingolipids in renal cell carcinoma tissues. J. Amer. Mass Spectrom. Soc. 28, 1562–1574 (2017).

27. Burla, B. et al. MS-based lipidomics of human blood plasma: a community-initiated position paper to develop accepted guidelines. J Lipid Res. 59, 2001–2017 (2018).

28. Triebl, A. et al. Shared reference materials harmonize lipidomics across MS based detection platforms and laboratories. J. Lipid Res. 61, 105e115 (2020).

29. Daemen, A. et al. Metabolite profiling stratifies pancreatic ductal adenocarcinomas into subtypes with distinct sensitivities to metabolic inhibitors. Proc. Natl. Acad. Sci. USA 112, E4410–E4417 (2015).

30. van der Hoeven, D. et al. Sphingomyelin Metabolism Is a Regulator of K-Ras Function. Mol Cell Biol. 38, e00373–17 (2018).

## References Methods

1. Wolrab, D., Chocholoušková, M., Jirásko, R., Peterka, O. & Holčapek, M. Validation of lipidomic analysis of human plasma and serum by supercritical fluid chromatography–mass spectrometry and hydrophilic interaction liquid chromatography–mass spectrometry. Analytical and Bioanalytical Chemistry 412, 2375–2388 (2020).

2. Liebisch, G., et al. Shorthand notation for lipid structures derived from mass spectrometry. Journal of Lipid Research 54, 1523–1530 (2013).

3. Fahy, E., et al. Update of the LIPID MAPS comprehensive classification system for lipids. Journal of Lipid Research 50, S9–S14 (2009).

4. Fahy, E., et al. A comprehensive classification system for lipids. Journal of Lipid Research 46, 839–862 (2005).

5. Rifai, N., Gillette, M.A., Carr, S.A. Protein biomarker discovery and validation: the long and uncertain path to clinical utility. Nature Biotechnology 24, 971–983 (2006).

6. Wolrab, D., et al. Determination of one year stability of lipid plasma profile and comparison of blood collection tubes using UHPSFC/MS and HILIC-UHPLC/MS. Analytica Chimica Acta 1137, 74–84 (2020).

7. Bligh, E.G. & Dyer, W.J. A Rapid method of total lipid extraction and purification. Canadian Journal of Biochemistry and Physiology 37, 911–917 (1959).

8. Lísa, M., Cífková, E., Khalikova, M., Ovčačíková, M. & Holčapek, M. Lipidomic analysis of biological samples: Comparison of liquid chromatography, supercritical fluid chromatography and direct infusion mass spectrometry methods. Journal of Chromatography A 1525, 96–108 (2017).

9. Cífková, E., et al. Lipidomic differentiation between human kidney tumors and surrounding normal tissues using HILIC-HPLC/ESI–MS and multivariate data analysis. Journal of Chromatography B 1000, 14–21 (2015).

10. Liebisch, G., et al. High throughput quantification of cholesterol and cholesteryl ester by electrospray ionization tandem mass spectrometry (ESI-MS/MS). Biochimica et Biophysica Acta (BBA) - Molecular and Cell Biology of Lipids 1761, 121–128 (2006).

11. Liebisch, G., Lieser, B., Rathenberg, J., Drobnik, W. & Schmitz, G. High-throughput quantification of phosphatidylcholine and sphingomyelin by electrospray ionization tandem mass spectrometry coupled with isotope correction algorithm. Biochimica et Biophysica Acta (BBA) - Molecular and Cell Biology of Lipids 1686, 108–117 (2004).

12. Liebisch, G., Drobnik, W., Lieser, B. & Schmitz, G. High-Throughput Quantification of Lysophosphatidylcholine by Electrospray Ionization Tandem Mass Spectrometry. Clinical Chemistry 48, 2217–2224 (2002).

13. Matyash, V., Liebisch, G., Kurzchalia, T.V., Shevchenko, A. & Schwudke, D. Lipid extraction by methyl-tert-butyl ether for high-throughput lipidomics. Journal of Lipid Research 49, 1137–1146 (2008).

14. Berry, K.A.Z. & Murphy, R.C. Electrospray ionization tandem mass spectrometry of glycerophosphoethanolamine plasmalogen phospholipids. Journal of the American Society for Mass Spectrometry 15, 1499–1508 (2004).

15. Liebisch, G., et al. Quantitative measurement of different ceramide species from crude cellular extracts by electrospray ionization tandem mass spectrometry (ESI-MS/MS). Journal of Lipid Research 40, 1539–1546 (1999).

16. Höring, M., Ejsing, C.S., Hermansson, M. & Liebisch, G. Quantification of Cholesterol and Cholesteryl Ester by Direct Flow Injection High-Resolution Fourier Transform Mass Spectrometry Utilizing Species-Specific Response Factors. Analytical Chemistry 91, 3459–3466 (2019).

17. Husen, P., et al. Analysis of Lipid Experiments (ALEX): A Software Framework for Analysis of High-Resolution Shotgun Lipidomics Data. PLOS ONE 8, e79736 (2013).

18. Chong, J. & Xia, J. MetaboAnalystR: an R package for flexible and reproducible analysis of metabolomics data. Bioinformatics 34, 4313–4314 (2018).

