## Supplementary tables guide for "Lipidomic profiling of human serum enables detection of pancreatic cancer"

**Overview of supplementary tables**

**Supplementary Table 1.** Concentrations of the internal standard (IS) mixture used for the lipidomic quantitation: **a,** Phase I (discovery) for UHPSFC/MS, shotgun MS, and MALDI-MS in lab 1. **b,** Phase II (qualification) and Phase III (verification) for UHPSFC/MS in lab 1. **c,** Phase II (qualification) for shotgun MS (LR and HR) in lab 2. **d,** Phase II (qualification) for RP-UHPLC/MS in lab 3.

**Supplementary Table 2.** Lipid shorthand nomenclature used throughout this work.

**Supplementary Table 3.** Molar concentrations of individual lipids measured in Phase I (discovery) together with the statistical evaluation. **a,** UHPSFC/MS. **b,** Shotgun MS. **c.** MALDI-MS.

**Supplementary Table 4.** Molar concentrations of individual lipids measured in Phase II (qualification) together with the statistical evaluation and normalized concentrations. **a,** UHPSFC/MS. **b,** Shotgun MS (LR). **c,** Shotgun MS (HR). **d,** RP-UHPLC/MS for molar concentrations before normalization. **e,** RP-UHPLC/MS for normalized concentrations after the transformation from lipid fatty acyl level to lipid species level.

**Supplementary Table 5.** Molar concentrations of individual lipids measured by UHPSFC/MS in Phase III (verification) together with the statistical evaluation and normalized concentrations.

**Supplementary Table 6.** Overview of studied human subjects in this study.

**Supplementary Table 7.** Clinical information for all samples used in this study.

**Supplementary Table 8.** Characteristics of OPLS-DA models for all MS based methods and all phases. **a,** Sensitivity, specificity, and accuracy values for training and validation sets. **b,** Summary of predictive components and orthogonal in X components.

**Supplementary Table 9.** Statistical parameters calculated from molar concentrations for the training set shown for all methods and all lipids with fold change ≥20%, p-value<0.05, and VIP value >1. **a,** Phase I (discovery). **b,** Phase II (qualification). **c,** Phase III (verification).

**Supplementary Table 10.** Predicted response values calculated from OPLS-DA models for all methods and all phases. **a,** Validation sets. **b,** Training sets.

**Supplementary Table 11.** Survival analysis data from Kaplan-Meier plots for individual methods in Phase II (qualification). **a,** Lipid species concentrations normalized to the NIST reference material, where values lower than median are classified as 0, and values higher than median are 1 for all lipids with p-value <0.05. **b,** Summary table of significant lipid species observed for multiple methods.

**Supplementary Table 12.** Multiple reaction monitoring (MRM) settings for RP-UHPLC/MS measurements in lab 3.
